# Prevalence and distribution of non-typhoidal *Salmonella enterica* serogroups and serovars isolated from normally sterile sites: a global systematic review

**DOI:** 10.1101/2023.06.13.23291315

**Authors:** Nienke N. Hagedoorn, Shruti Murthy, Megan Birkhold, Christian S. Marchello, John A. Crump, Vacc-iNTS Consortium

**Affiliations:** Centre for International Health, University of Otago, Dunedin, New Zealand; Department of Surgery, University of Maryland School of Medicine, Baltimore, Maryland, United States of America

**Author notes:** Corresponding author: Professor John A Crump, Centre for International Health, University of Otago PO Box 56, Dunedin 9054, New Zealand. Nienke N. Hagedoorn –, Shruti Murthy –, Megan Birkhold –, Christian S. Marchello -, John A. Crump.

**Keywords:** Non-typhoidal *Salmonella*, systematic review, meta-analysis, prevalence, *Salmonella* vaccines

## Abstract

To inform vaccine development strategies, we aimed to systematically review evidence on the prevalence and distribution of non-typhoidal *Salmonella enterica* serogroups and serovars. We searched four databases from inception through 4 June 2021. We included articles that reported at least one non-typhoidal *Salmonella enterica* strain by serogroup or serovar isolated from a normally sterile site. Of serogrouped isolates, we pooled the prevalence of serogroup O:4, serogroup O:9, and other serogroups using random-effects meta-analyses. Of serotyped isolates, we pooled the prevalence of *Salmonella* Typhimurium (member of serogroup O:4), *Salmonella* Enteritidis (member of serogroup O:9), and other serovars. Of 82 studies yielding 24,258 serogrouped isolates, pooled prevalence (95% CI) was 44.7% (36.3%-48.3%) for serogroup O:4, 45.4% (36.9%-49.0%) for serogroup O:9, and 9.9% (6.1%-13.3%) for other serogroups. Pooled prevalence (95%CI) was 36.8% (29.9%-44.0%) for *Salmonella* Typhimurium, 37.8% (33.2%-42.4%) for *Salmonella* Enteritidis, and 18.4% (11.4%-22.9%) for other serovars. Of global serogrouped non-typhoidal *Salmonella* isolates from normally sterile sites, serogroup O:4 and O:9 together accounted for 90%. Of global serotyped isolates, serovars Typhimurium and Enteritidis together accounted for 75%. Vaccine development strategies covering serogroups O:4 and O:9, or serovars Typhimurium and Enteritidis, have the potential to prevent the majority of non-typhoidal *Salmonella* invasive disease.

PROSPERO:CRD42022376658.

**Key results:**

- Invasive infections in normally sterile sites caused by non-typhoidal *Salmonella* have a case fatality ratio of 15%.
- Vaccine products for human non-typhoidal *Salmonella* are in currently in development.
- For future vaccine strategies, we provide global non-typhoidal *Salmonella* serogroup and serovar coverage by region and by age groups.
- Serogroups O:4 and O:9 account for 90% of isolates of non-typhoidal *Salmonella enterica* from normally sterile sites.
- Serovars Typhimurium and Enteritidis, members of serogroup O:4 and O:9, respectively, cover 75% of isolates of non-typhoidal *Salmonella enterica* from normally sterile sites.

## Introduction

Worldwide, bacterial infections caused by *Salmonella enterica* are responsible for substantial burden of illness and death.^1^ Invasive *Salmonella* disease can be grouped into that caused by the typhoidal *Salmonella* serovars Typhi, Paratyphi A, Paratyphi B, and Paratyphi C, and that caused by the non-typhoidal *Salmonella* serovars.^2^ Non-typhoidal *Salmonella* (NTS) is classified into 67 serogroups based on the O antigen, the polysaccharide component of the lipopolysaccharide of the outer membrane, and can be further classified into more than 2,500 serovars based on the flagellar H antigen, and a virulence-related capsular polysaccharide antigen, Vi.^3^

NTS serovars usually have their reservoirs in animals. Humans are often infected by the consumption of contaminated foods of animal origin through contaminated water, or by the faecal-oral route during contact with reservoir species.^4–6^ However, it has been suggested humans could be a reservoir of NTS serovars and sequence types (STs) associated with invasive disease in Africa.^7^ ^8^ NTS may cause diarrheal disease that is generally self-limiting in healthy adults, whereas invasive disease can develop in the absence of current or recent diarrhoea and carries a case fatality ratio of 15%. ^9^ Persons at increased risk for invasive disease include those with human immunodeficiency virus (HIV) infection, current or recent malaria, and children with malnutrition.^1 10 11^ We recently performed a global systematic review on the complications and mortality of NTS invasive disease.^9^ We reported that *Salmonella* Typhimurium and *Salmonella* Enteritidis members of serogroups O:4 and O:9, respectively, were the most abundant serotypes collectively accounting for 78% to 94% of isolates.^9^ ^12^ In addition, specific STs including *Salmonella* Typhimurium ST313 and *Salmonella* Enteritidis ST11 are prevalent and are associated with antimicrobial resistant infections.^13–16^

Vaccine candidates for NTS invasive disease in preclinical or early clinical phase of development have been described in detail by Baliban *et al* (see Table 1).^17–24^ In order to assess the potential serogroup and serovar coverage of vaccine candidates and future candidates, we extended our previous systematic review to estimate the prevalence and distribution of non-typhoidal *Salmonella enterica* serogroups and serovars isolated from normally sterile sites.

**Table 1–.** Non-typhoidal *Salmonella enterica* invasive disease vaccines in development and estimated isolate coverage

| Type of vaccine | Vaccine antigen<br>Serovar (serogroup) | Estimated isolate coverage in non-typhoidal *Salmonella* based on estimated prevalence |  |
| --- | --- | --- | --- |
|  |  | Serogroup: assuming cross-protection for serovars within the same serogroup | Serovar: coverage at the serovar level without assuming any cross-protection |
| Generalised Modules for Membrane Antigens (GMMA) |  |  |  |
| *Salmonella* Typhimurium GMMA, as part of a 2-component *Salmonella* Enteritidis/*Salmonella* Typhimurium GMMA vaccine <sup>23</sup> | *Salmonella* Enteritidis (O:9) | 30.4% | 27.4% |
|  | *Salmonella* Typhimurium (O:4) | 63.3% | 59.7% |
| Glycoconjugates |  |  |  |
| Non-typhoidal *Salmonella* Core and O-polysaccharide conjugated to the flagellin protein.<br>Trivalent *Salmonella* Conjugate Vaccine <sup>20-22</sup> | *Salmonella* Enteritidis (O:9) | 30.4% | 27.4% |
|  | *Salmonella* Typhimurium (O:4) | 63.3% | 59.7% |
|  | *Salmonella* Typhi Vi (O:9)* | _* | _* |
| Live-attenuated approaches |  |  |  |
| *Salmonella* Typhimurium strain LH1160 chicken isolate <sup>18</sup> | *Salmonella* Typhimurium (O:4) | 63.3% | 59.7% |
| Human *Salmonella* Typhimurium enterocolitis isolate <sup>19</sup> | *Salmonella* Typhimurium (O:4) | 63.3% | 27.4% |

<sup>^</sup> Adapted from Baliban et al. <sup>17 24</sup>
\* *Salmonella* Typhi not part of this review

## Methods

### Study design, selection criteria and search strategy

Our study is reported according to the Preferred Reporting Items for Systematic Reviews and Meta Analyses (PRISMA) model.^25^ Our study was of published data and therefore institutional review board approval was not required. The study protocol was registered in Prospective Register of Systematic Reviews (PROSPERO) at 27 November 2022 and the study protocol is available online (CRD42022376658).^26^

Our study was an extension of the previous global systematic review and meta-analysis described by Marchello *et al.* that reported complications and the case fatality ratio among persons with NTS invasive disease.^9^ In brief, a literature search was performed in Embase, MEDLINE, Web of Science, and PubMed from database inception through 4 June 2021 with key words including non-typhoidal *Salmonella*; specific serovars such as *Salmonella* Typhimurium, and *Salmonella* Enteritidis; and mortality or complications.^9^ We did not restrict the search by language, country, or date. We included primary research articles that were peer-reviewed and reported the number of isolates of NTS and at least one NTS strain by serogroup or serovar, confirmed by culture of samples taken from normally sterile sites (e.g., blood, bone marrow). We excluded articles of specific disease groups (e.g. severe malaria), that focused on one specific serovar, or that focused on antimicrobial resistant isolates. In addition, we excluded case reports, case series, policy reports, commentaries, and conference abstracts. In contrast to the original systematic review, studies for this extension were eligible when they reported the number of at least one NTS strain by serogroup or serovar irrespective of whether they reported on the proportion of complications or deaths.

Search results from each electronic database were downloaded, imported into Endnote X20 (Clarivate, London, United Kingdom), and duplicates were subsequently removed. To assess eligibility for the present study, the 291 articles that passed the initial title and abstract review were reviewed again by NNH and SM. Final decision on eligibility was resolved through discussion or by decision of a third reviewer (JAC).

### Data abstraction

Data were abstracted in Google Forms (Google LLC, USA). Abstracted data included study characteristics such as country, data collection period, United Nations (UN) region,^27^ and the age groups children ≤15 years, adults >15 years, or mixed ages. We recorded the total of number of isolates of NTS, and the number of isolates for each reported serogroup, or each reported serovar. We classified isolates as ‘not further identified’ when reported as a mixed group of unspecified serogroups or serovars, or when isolates were classified as NTS and were not reported to the serogroup or serovar level. We could not reliably abstract the number of isolates of NTS that were not serogrouped or serotyped. Serovars were grouped into serogroups following the antigenic formulae of *Salmonella* serovars (Supplemental file S1).^3^ We reported serogroups according to the current serogroup designation with the historic designation provided in parenthesis at first use. For example, serogroup O:4 (B). We combined serogroups with <10 isolates in ‘other serogroups with <10 isolates’. Serovars that could not be serogrouped were classified as undesignated. We sought to abstract data on serogroup or serovar by host risk factor data, and to analyse by host risk factor when reported sufficiently frequently to make that possible. Next, we recorded the reported serotyping methods including the agglutination test, polymerase chain reaction, multilocus sequence typing, other, or unclear. For articles that reported that ‘standard methods’ were used, we assumed that the agglutination test was used. For those that used agglutination tests, we recorded whether the anti-serum panel available to the laboratory was reported.

#### Bias assessment

Risk of bias was assessed using the same methodology as the primary article^9^ (Supplemental File S2).^28–30^ We evaluated study design, study setting, patient selection, and definitions for microbiology methods. Risk of bias was assessed independently by one of CSM, MB, NNH, or SM, and each question was scored as unknown, low, or high risk. The scores were aggregated to assign each study as having a low, moderate, or high risk of bias. Conflicts were resolved through discussion or by decision of a third author.

### Data analysis

First, we described the characteristics of the included articles. Second, we reported the proportion of serogroups and for serovars. Third, we explored longitudinal trends by grouping the articles by decade according to the median year of the reported period of data collection. Fourth, we assessed the geographic distribution of serogroups and serovars of all NTS isolates by UN region and their rank order in prevalence. Fifth, we described the distribution of serogroups and serovars by HIV infection status of participants. We assumed that only one isolate was reported per participant. We compared the proportion of serogroup O:4, serogroup O:9 and other serogroups, and the proportion of *Salmonella* Typhimurium, *Salmonella* Enteritidis and other serovars, according to HIV infection status using the Chi^2^ test.

#### Meta-analysis

For isolates identified to the serogroup level, we pooled the prevalence across serogroup O:4, serogroup O:9, and other serogroups. We performed a meta-analysis in MetaXL version 5.3 (EpiGear International Pty Ltd., Australia) using the DerSimonian-Laird random-effects model for multiple categories with the double arcsine transformation.^31^ We performed subgroup analyses by UN region, and by age group. In addition, for isolates identified to the serovar level, we pooled the prevalence across *Salmonella* Typhimurium, *Salmonella* Enteritidis, and other serovars. Heterogeneity across studies was assessed using forest plots, Chi^2^ test, *I*^2^ statistic and Tau2 (τ^2^). We considered an I^2^ value of 0‒49% as low, 50‒74% as moderate, and ≥75% as substantial heterogeneity.^32^ We considered a p-value <0.05 as indicating significant heterogeneity.

In addition, we estimated the proportion of NTS isolates that might be covered by current vaccines in development based on the prevalence of serogroups and serovars (Table 1). While the mechanism and immune correlates of protection of vaccines in development against NTS invasive disease are not known, we examined potential coverage at both the serogroup and serovar level. For potential coverage at the serogroup level, we looked at coverage assuming that vaccines would provide cross-protection for serovars within the same serogroup. We also explored coverage with the addition of other common serogroups and serovars. Lastly, we performed a sensitivity analysis repeating the meta-analyses while excluding one article that provided more than a third of the isolates. Apart from the meta-analysis that was performed in MetaXL, other analyses were performed in R version 4.2 (packages: dplyr, forestploter, ggplot2, tidyr).

## Results

### Article characteristics

Of the 291 full-text articles, 82 articles were included (Supplemental Figure S1; Supplemental File S3). Of the 82 articles, 31 (37.8%) reported data from Africa, 21 (25.6%) from Asia, 20 (24.4%) from Europe, 9 (10.9%) from the Americas, and 1 (1.2%) from Oceania (Supplemental File S4, Supplemental Figure S2). UN subregions lacking data included Northern Africa, Central Asia, Micronesia, Melanesia, and Polynesia. Data reported by the included articles were collected from 1941 through 2019 with a median (IQR) duration of data collection of 6 (3-10) years. The majority of articles collected data from a hospital setting (n=71, 86.6%). The population was of mixed children and adults in 36 (43.9%) articles, children in 33 (40.2%) articles, and adults in 13 (15.8%) articles.

Overall, 61 (74.4%) articles were assessed as high risk for bias, 21 articles (25.6%) as moderate risk, and no articles (0.0%) were low risk (Supplemental Figure S3). Domains with the highest proportion of high risk for bias included study setting (n=70, 85.4%), and microbiology methods (n=61, 74.4%). Reported methods for serotyping were agglutination testing for 49 (59.8%) articles, combination of methods for 5 (6.1%), multilocus sequence typing for 1 (1.2%), and unclear for 27 (32.9%). Of the 49 articles using agglutination testing, 17 articles reported details on the anti-serum panel available.

### NTS serogroups and serovars

The number of NTS isolates ranged from 1 to 10,139 per study with a median (IQR) of 44 (16-101) isolates per study, yielding a total of 26,280 isolates. Of the 26,280 isolates, 15,348 (58.5%) were classified as serogroup O:4 (B), 7,386 (28.2%) as serogroup O:9 (D1), 1,065 (4.1%) as serogroup O:7 (C1), 250 (1.0%) as group O:8 (C2-C3), 2,018 isolates (7.7%) were not further identified (Figure 1A).

**Figure 1–.**
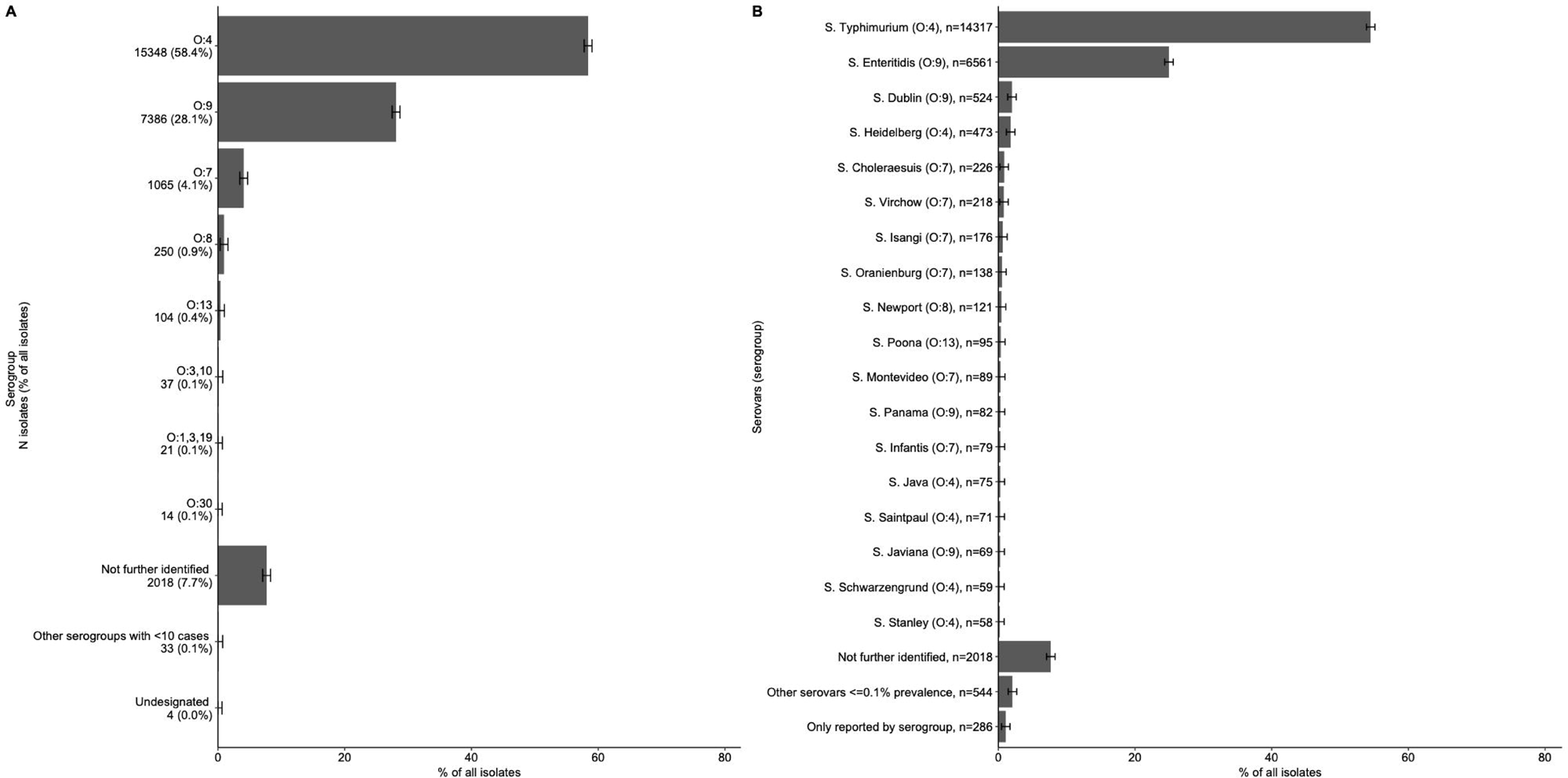
Proportion of non-typhoidal *Salmonella enterica* serogroups (A) and serovars (B) isolated from normally sterile sites, global, 1941-2019 (n=26,280) Legend: The error bars represent the 95%CI of the proportion. The proportion of each serovar by serogroup is provided in Supplemental File S5. A: Other serogroups with <10 cases: O:2 (A), n=1; O:9,46 (D2), n=1; O:11 (F), n=8; O:16 (I), n=4; O:18 (K), n=5; O:21 (L), n=3; O:28 (M), n=3; O:35 (O), n=3; O:38 (P), n=1; O:40 (R), n=4. B: The list of other serovars <=0.1% is provided in Supplemental File S6.

Of the 26,280 isolates, 14,317 (54.5%) isolates were serotyped as *Salmonella* Typhimurium, 6,561 (25.0) as *Salmonella* Enteritidis, 524 (2.0%) as *Salmonella* Dublin, 473 (1.8%) as *Salmonella* Heidelberg, 226 (0.9%) as *Salmonella* Choleraesuis, and 218 (0.8%) as *Salmonella* Virchow (Figure 1B). The serogroup distribution across decades is presented in Supplemental Figure S4.

### Geographic distribution of serogroups and serovars

Of 24,258 serogrouped isolates, 17,350 (71.5%) were from the African region, 3,173 (13.1%) from Europe, 2,645 (10.9%) from the Americas, 1,087 (4.5%) from Asia, and 3 (<0.1%) from Oceania. For Africa, Europe, the Americas, and Asia, serogroup O:4 and serogroup O:9 were the two most common serogroups (Supplemental Figure S5). In Oceania, the three isolates were serogroup O:7. Serogroup O:4 was the most common serogroup in 12,705 (73.2%) of 17,350 isolates from the African region and in 1,336 (50.5%) of 2,645 isolates from the Americas. Serogroup O:9 accounted for 1,810 (57.0%) of 3,173 isolates in the European region. Of 1,087 isolates in Asia, 423 (38.9%) were serogroup O:4 and 397 (36.5%) were serogroup O:9. Serogroup O:7 was the third ranked serogroup in Africa, Europe, the Americas and Asia.

For Africa, Europe, the Americas, and Asia, *Salmonella* Typhimurium and *Salmonella* Enteritidis were the two most common serovars (Figure 2). In Oceania, the three isolates were *Salmonella* Virchow. *Salmonella* Typhimurium accounted for 12,684 (73.1%) of 17,351 isolates from the African region and 705 (26.6%) of 2,645 isolates from the Americas. *Salmonella* Enteritidis accounted for 1,518 (48.9%) of 3,103 isolates from European region. Of 874 serotyped isolates from the Asian region, *Salmonella* Enteritidis accounted for 318 (36.4%) and *Salmonella* Typhimurium for 280 (32.0%). Variation was observed for the serovars that ranked third, fourth and fifth. The third ranked serovar was *Salmonella* Dublin in Africa and Europe, *Salmonella* Heidelberg in the Americas, and *Salmonella* Choleraesuis in Asia. In addition, variation was observed for the proportion of serovars with a ranking of sixth and higher across UN regions: 786 (29.7%) of 17,351 in the Americas, 508 (16.4%) of 3,103 in Europe, 86 (9.8%) of 874 in Asia, and 51 (0.3%) of 13,751 in Africa.

**Figure 2–.**
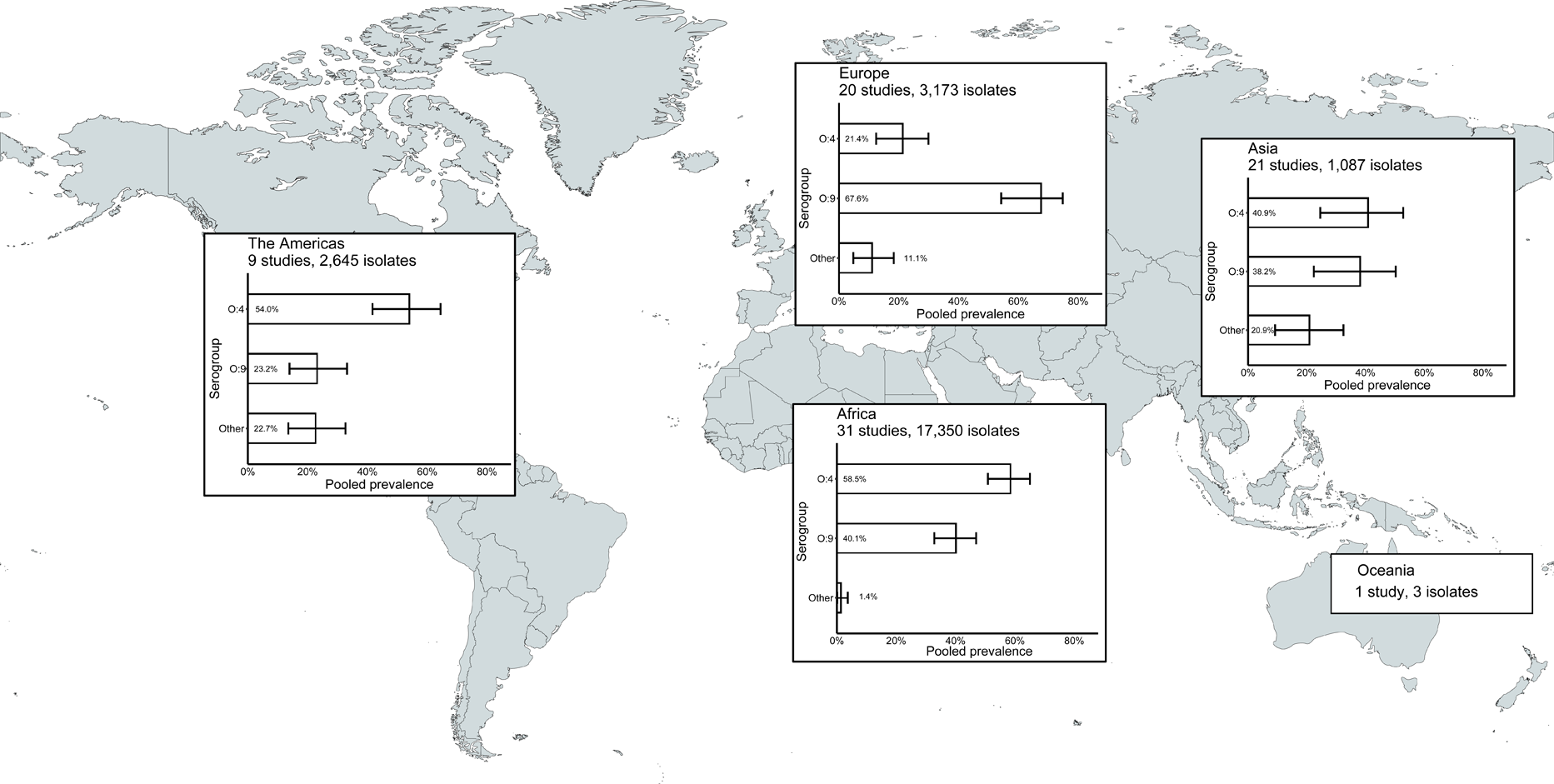
Prevalence of non-typhoidal *Salmonella enterica* serovars from normally sterile sites ranked among top five, by UN region, 1941-2019 (79 articles, 23,976 isolates) Legend: The error bars indicate 95%CI of the prevalence. *Others include other serovars that had rank sixth or higher. Data for Oceania is described in the Results. Global map was downloaded from mapchart.net.

### *Salmonella* serogroup and serovar by HIV-infection status

Of the 2,000 serogrouped isolates from ten articles with available HIV infection status, 1,743 (87.2%) were from HIV-infected participants (Table 2). The prevalence of serogroup O:4 in 1,174 (67.4%) of 1,743 among isolates from HIV-infected participants was higher than the 89 (34.6%) of 257 among isolates from HIV-uninfected participants (p<0.01). The prevalence of serogroup O:9 in 559 (32.1%) of 1,743 among isolates from HIV-infected participants was lower than 157 (61.1%) of 257 among isolates from HIV-uninfected participants (p<0.01). The prevalence of serogroups other than O:4 or O:9 is presented in Table 2. The prevalence of *Salmonella* Typhimurium was 1,174 (67.4%) of 1,743 among isolates HIV-infected participants and 88 (34.1%) of 258 among isolates from HIV-uninfected participants (p<0.01). The prevalence of *Salmonella* Enteritidis was 493 (28.3%) of 1,743 isolates among HIV-infected participants and 147 (56.9%) of 258 among isolates from HIV-uninfected participants (p<0.01). The prevalence of other serovars is presented in table 2.

**Table 2–.**
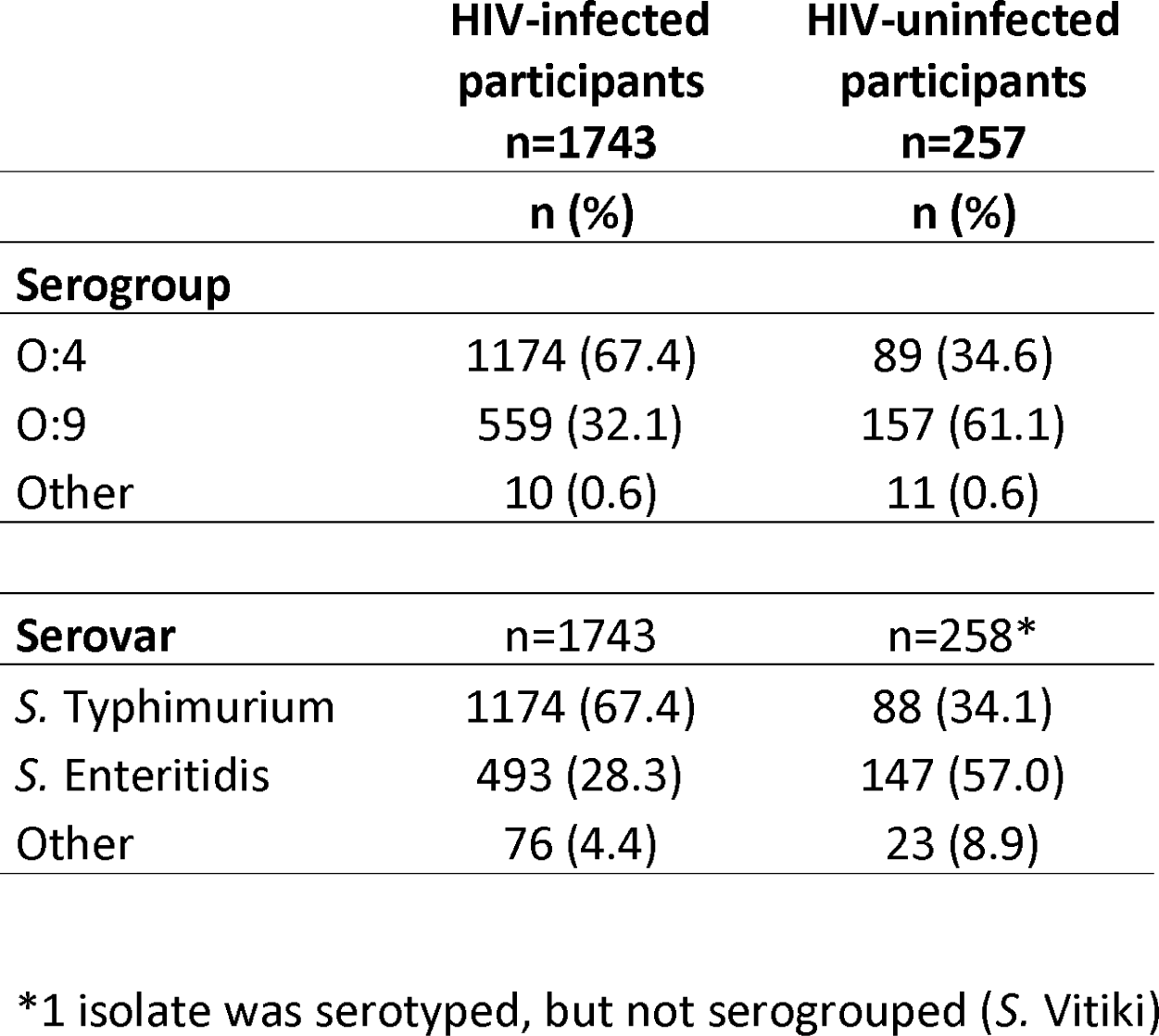
The proportion of non-typhoidal *Salmonella enterica* serogroups and serovars by HIV infection status of participants, isolated from normally sterile sites, global, 1941-2019 (10 articles, 2,000 serogrouped isolates)

### Overall pooled prevalence of NTS serogroup, by UN region, and by age

In the meta-analysis of the 82 studies with 24,258 serogrouped isolates, pooled prevalence (95%CI) was 44.7% (36.3%-48.3%) for group O:4, 45.4% (36.9%-49.0%) for group O:9, and 9.9% (6.1%-13.3%) for other serogroups (I^2^= 98.5%, p<0.001) (Supplemental Figure S6). For subgroup analysis by UN region, the pooled prevalence of group O:4 was 58.5% (50.9%-65.0%) in Africa (31 studies, 17,350 isolates, I^2^= 98.1%, p<0.001), 54.0% (41.7%-64.4%) in the Americas (9 studies, 2,645 isolates, I ^2^= 74.4%, p<0.001), 40.9% (24.6%-52.8%) in Asia (21 studies, 1,187 isolates, I^2^= 95.4%, p<0.001), and 21.4% (12.4%-29.9%) in Europe (20 studies, 3,244 isolates, I^2^= 95.3%, p<0.001) (Figure 3, Supplemental Figure S7). For group O:9, the pooled prevalence was 40.1% (32.8%-46.9%) in Africa, 23.2% (13.9%-33.2%) in Americas, 38.2% (22.4%-50.3%) in Asia, and 67.6% (54.2%-74.8%) in Europe. For other serogroups, the pooled prevalence was 1.4% (0.1%-3.7%) in Africa, 22.7% (13.5%-32.6%) in the Americas, 20.9% (9.2%-32.5%) in Asia, and 11.1% (4.8%-18.3%) in Europe. A meta-analysis could not be performed as only one study reporting three isolates included data from Oceania.

**Figure 3–.**
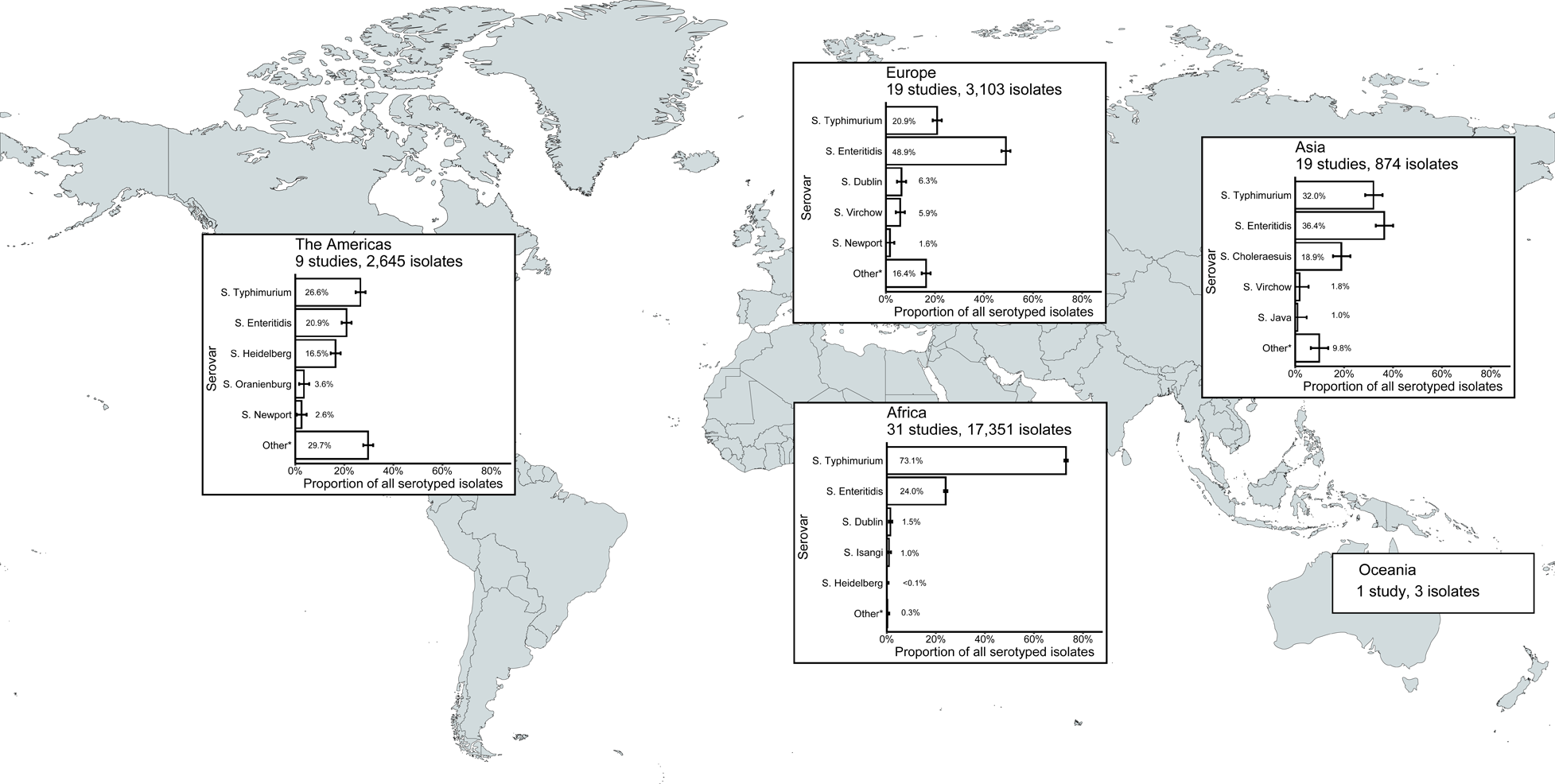
Meta-analysis of prevalence of non-typhoidal *Salmonella enterica* from normally sterile sites for serogroup O:4, serogroup O:9, and other serogroups by UN region, 1941-2019 (82 articles, 24,258 isolates) Legend: The error bars indicate 95%CI of pooled prevalence. Data for Oceania was not pooled and is described in the Results. Global map was downloaded from mapchart.net.

Pooled prevalence (95% CI) of serogroup O:4 was 37.9% (26.5%-44.3%) in mixed ages (36 studies, I ^2^= 99.2%, p<0.001), 51.0% (39.8%-57.8%) in children (33 studies, I^2^= 95.2%, p<0.001), and 48.3% (28.1%-65.1%) in adults (13 studies, I^2^= 92.5%, p<0.001) (Supplemental Figure S8). The pooled prevalence of serogroup O:9 was 48.0% (35.4%-53.9%) in mixed ages, 43.0% (32.4%-50.1%) in children, 44.1% (24.6%-61.2%) in adults. The pooled prevalence of other serogroups was 14.0% (7.3%-20.0%) in mixed ages, 6.0% (2.1%-10.8%) in children, and 7.5% (0.2%-20.4%) in adults.

### Pooled prevalence of NTS serovars

In meta-analysis of isolates reported to the serovar level, the pooled prevalence (95%CI) was 36.8% (29.9%-44.0%) for *Salmonella* Typhimurium, 37.8% (33.3%-42.4%) for *Salmonella* Enteritidis, and was 18.4% (11.4%-22.9%) for other serovars (79 studies, 23,976 isolates, I^2^= 99.1%, p<0.001) (Supplemental Figure S9).

### Estimated coverage for vaccine products in development

Assuming cross-protection for serovars within the same serogroup, the estimated coverage of vaccines products was 63.3% in serogroup O:4 and was 30.4% in serogroup O:9. For serogroups O:4 and O:9 combined, the estimated coverage was 93.7% for isolates classified to the serogroup level (Table 1). The addition of serogroup O:7 and serogroup O:8 would increase the estimated coverage with 4.4% and 1.0%, respectively. For serovars, the estimated coverage was 59.7% for vaccines targeting *Salmonella* Typhimurium and was 27.4% for vaccines targeting *Salmonella* Enteritidis. For Salmonella Typhimurium and Salmonella Enteritidis together, the estimated coverage was 87.1%. The addition of *Salmonella* Dublin, *Salmonella* Heidelberg, or *Salmonella* Choleraesuis would increase the estimated coverage with 2.2%, 1.9%, and 0.9%, respectively.

The sensitivity analysis excluding the one article that provided 10,139 (38.6%) of all 26,280 isolates yielded similar results (not shown).^33^

## Discussion

Our global systematic review and meta-analysis of the prevalence and geographic distribution of non-typhoidal *Salmonella enterica* serogroups and serovars isolated from normally sterile sites showed that the prevalence of serogroup O:4 and serogroup O:9 of all grouped isolates was estimated to be 44.7% and 45.3%, respectively. We estimated the prevalence of *Salmonella* Typhimurium, a member of serogroup O:4, and *Salmonella* Enteritidis, a member of serogroup O:9, to be 36.8% and 37.8%, respectively. Our study identified some regional variation in serogroup and serovar distribution. Serogroup O:4 accounted for the largest proportion of isolates in Africa, driven by the high prevalence of *Salmonella* Typhimurium, and serogroup O:9 accounted for the largest proportion of isolates in Europe, driven by both *Salmonella* Enteritidis and *Salmonella* Dublin.

Our data align with a previous systematic review through 2014 that focused on Africa and reported that *Salmonella* Typhimurium and *Salmonella* Enteritidis were the most prevalent serovars causing NTS invasive disease, accounting for more than 90% of isolates.^10^ Of articles included in our systematic review, regions that provided 20 or more studies included the UN regions of Africa, Asia, and Europe with the majority of NTS isolates originating from the UN African region. Data from UN region Oceania was limited to one article that yielded just three isolates. Notably, UN subregions lacking data included Northern Africa, Central Asia, Micronesia, Melanesia, and Polynesia. (Supplemental Figure S2). In some countries, the lack of data could be partially explained by the lower incidence of NTS disease.^1^ Additionally, high-income countries are more likely to report data on NTS invasive disease in routine surveillance systems reports that were out of scope for this study. Another contributor to limited data for some regions included that *Salmonella* isolates were not always reported to the serogroup or serovar level. It is likely that for some countries NTS invasive disease was prevalent, but data on serogroup and serovar was not available due to lack of strain typing.^1^

We showed that serogroup O:4, driven by *Salmonella* Typhimurium, was more often isolated from HIV-infected persons and that serogroup O:9 was more often isolated from HIV-uninfected participants. It may be that *Salmonella* Typhimurium, the most common serovar in serogroup O:4, could have distinct pathogenesis in HIV-infected persons. Previous *in vitro* studies on the pathogenesis of *Salmonella* Typhimurium ST313 in HIV-infected and HIV-uninfected individuals have shown that serum bactericidal activity is lower in HIV-infected persons compared with those without HIV.^34^ ^35^ Future efforts to reduce the burden of NTS invasive disease among those with HIV infection should focus on both preventing exposure to reservoirs and sources of the pathogen, and also on improving access to antiretroviral therapy to aid in restoring the immune function. In addition, future NTS vaccine trials need to assess the immune response and level of protection among HIV-infected individuals as well as other groups at greater risk for disease.

Our meta-analysis showed there was substantial heterogeneity across included articles. This was expected as we included articles from a range of UN regions and settings including both hospital-based and community-based studies. In addition, the large proportion of articles that were classified as high-risk of bias could have contributed to the heterogeneity. The majority of the included articles were assessed to be at a high risk of bias for microbiology primarily due to incomplete reporting of microbiological methods. In addition, for one third of articles, the method used for *Salmonella* serotyping was unclear. We suggest that reporting of microbiology methods should be improved. This should include details of the serotyping methods used and any external quality assurance procedures. When agglutination testing is used, the details of the battery of anti-sera available in the testing laboratory should be reported. For *in silico* serotyping reporting the bioinformatics pipeline and pipeline version used would assist with interpretation.

For vaccine products currently in development, we provided estimates for potential coverage based on estimated prevalence of serogroup and serovar. Assuming that vaccines would provide cross-protection for serovars in the same serogroup, live-attenuated *Salmonella* Typhimurium vaccine products would have the potential to cover 63.3% of all serogrouped isolates. If no within serogroup cross-protection is assumed, the target of *Salmonella* Typhimurium would cover 59.7% of all serotyped isolates. Assuming within serogroup cross-protection, the *Salmonella* Typhimurium and *Salmonella* Enteritidis Generalised Modules for Membrane Antigens (GMMA) vaccine and the trivalent *Salmonella* glycoconjugate vaccine targeting both *Salmonella* Typhimurium and *Salmonella* Enteritidis have the potential to cover 94% of all isolates classified to the serogroup level. The addition of coverage for serogroup O:7 will increase the coverage with 4%. If no within serogroup cross-protection is assumed, vaccines targeting *Salmonella* Typhimurium and *Salmonella* Enteritidis alone would cover approximately 87% of isolates. Further addition of *Salmonella* Dublin or *Salmonella* Heidelberg would increase the coverage with 2%. Since vaccines vary considerably in the range of antigens used and in likely correlates of protection, assumptions about coverage and cross-protection should be interpreted with caution. In addition, our coverage estimates would vary by region since we identified regional variation of serogroups and serovars.

In the Democratic Republic of the Congo, the emergence of a *Salmonella* Typhimurium O:5-negative variant has been reported which is lacking O:5 specificity.^36^ ^37^ This O:5-negative variant represented 37% of all *Salmonella* Typhimurium in the study by Tack *et al.* For vaccine products based on O:4 antigens, such as the *Salmonella* glycoconjugate vaccine, the emergence of this O:5-negative variant may not influence their potential coverage. Nevertheless, the fact that O antigens could be lost over time poses a risk to vaccine coverage.^38^

We searched multiple databases without restriction on time period, language, country, or the number of isolates. To our knowledge, our study is the first to provide global prevalence data for invasive NTS by serogroup and serovar by region. However, our study has some limitations. First, we based our study on a previous publication by our research group that focused on complications and mortality of NTS invasive disease. Therefore, the search strategy included the search terms ‘complications’ and ‘mortality’ that were not necessarily in scope for the present study and may have resulted in missing articles relevant to our research question. To be as comprehensive as possible, all articles that passed title and abstract review for the initial study were re-screened for eligibility for the present study. Second, we acknowledge that 7.7% of our isolates were not further classified in our review. The group of isolates not further classified included both isolates that were typed to the serogroup or serovar level and were reported as ‘other’ by the original authors, as well as isolates that were not typed. We did not request additional data from the authors of eligible papers. To ensure that our estimates were based on known serogroups and serovars, we excluded isolates that were not further classified to the serogroup or serovar level. As such, our study might overestimate the prevalence of serogroups or serovars for which antisera are more commonly available. For instance, in the African region the proportion of serovars with ranking sixth and higher was <1%, suggesting that the serovars ranked first through fifth were the most dominant. Also, in Africa the range of antisera might have been more limited than in other regions.^39^ ^40^ Third, we focused our analysis to the prevalence of serogroups and serovars and did not consider sequence types such as *Salmonella* Typhimurium ST313 and *Salmonella* Enteritidis ST11. While this likely has limited relevance for vaccine development, we were unable to evaluate the distribution of major sequence types. Fourth, while we appreciate the importance of host risk factors other than HIV, such as malnutrition and malaria, for NTS invasive disease, these host risk factors were not reported sufficiently frequently to be evaluated in our study. ^33^

## Conclusion

Of global serogrouped NTS from normally sterile sites, serogroup O:4 and O:9 accounted for 90% isolates, and of global serotyped NTS from normally sterile sites *Salmonella* Typhimurium and Enteritidis accounted for 75% of isolates. We observed some geographical variation in serogroup and serovar distribution. Among UN regions, serogroup O:4 had the highest prevalence in the African region, driven by *Salmonella* Typhimurium, and serogroup O:9 had the highest prevalence in the European region, driven by *Salmonella* Enteritidis and *Salmonella* Dublin. Vaccine development strategies that cover both serogroups O:4 and O:9, or *Salmonella* Typhimurium and *Salmonella* Enteritidis, have the potential to prevent the majority of NTS invasive disease.

## Data Availability

Data underlying this study will be made available upon acceptance of publication in a repository.

## Abbreviations

GMMA: Generalized Modules for Membrane Antigens
NTS: non-typhoidal *Salmonella*
PROSPERO: Prospective Register of Systematic Reviews
UN: United Nations

## Acknowledgements

**Vacc-iNTS Consortium Collaborators**

Francis Agyapong (Kwame Nkrumah University of Science and Technology Kumasi); Francesco Berlanda Scorza (GSK Vaccines Institute for Global Health); Gianluca Breghi (Fondazione Achille Sclavo); Rocío Canals (GSK Vaccines Institute for Global Health); Fabio Fiorino (University of Siena); Melita A Gordon (University of Liverpool); Jong-Hoon Kim (International Vaccine Institute); Brama Hanumunthadu (University of Oxford); Jan Jacobs (Institute of Tropical Medicine Antwerp); Samuel Kariuki (Kenya Medical Research Institute); Stefano Malvolti (MM Global Health Consulting); Carsten Mantel (MM Global Health Consulting); Florian Marks (University of Cambridge and International Vaccine Institute); Donata Medaglini (Università di Siena and Sclavo Vaccines Association); Esther Muthumbi (KEMRI-Wellcome Trust Research Programme); Tonney S. Nyirenda (University of Malawi); Robert Onsare (Kenya Medical Research Institute); Ellis Owusu-Dabo (Kwame Nkrumah University of Science and Technology Kumasi); Elena Pettini (University of Siena); Maheshi Ramasamy (University of Oxford); J. Anthony Scott (KEMRI-Wellcome Trust Research Programme); Bassiahi Abdramane Soura (University of Ouagadougou); Tiziana Spadafina (Sclavo Vaccines Association); Bieke Tack (Institute of Tropical Medicine Antwerp)

## Captions supplementary material

**Supplemental File S1** – Serogroup classification based on serogroups and serovars identified in the systematic review, global, 1941-2019 [3]

**Supplemental File S2** – Bias assessment methods, adopted from Marchello, 2022 ^9^

**Supplemental File S3** - Details of included articles identified in the global systematic review on prevalence of serogroups and serovars of non-typhoidal *Salmonella enterica* isolated from normally sterile sites, 1941 to 2019 (82 articles)

**Supplemental File S4** – Descriptive characteristics by article and by isolates identified in the global systematic review on prevalence of serogroups and serovars of non-typhoidal *Salmonella enterica* isolated from normally sterile sites, 1941 to 2019

**Supplemental File S5** - The proportion of non-typhoidal *Salmonella enterica* isolates by serovar for all isolates and the proportion of each serovar by serogroup identified in the global systematic review on prevalence of serogroups and serovars of non-typhoidal *Salmonella enterica* from normally sterile sites, 1941 to 2019 (26,280 isolates)

**Supplemental File S6** ‒ Serovars with prevalence ≤0.1% identified in the global systematic review on prevalence of serogroups and serovars of non-typhoidal *Salmonella enterica* isolated from normally sterile sites, 1941 to 2019 (26,280 isolates)

**Supplemental Figure S1** – PRISMA flowchart of study selection process for the global systematic review on prevalence of serogroups and serovars of non-typhoidal *Salmonella enterica* from normally sterile sites, 1941 to 2019

**Supplemental Figure S2** - Global distribution of number of articles per country identified in the global systematic review on prevalence of serogroups and serovars of non-typhoidal *Salmonella enterica* isolated from normally sterile sites, 1941 to 2019 (82 articles)

**Supplemental Figure S3** – Bias assessment of the global systematic review on prevalence of serogroups and serovars of non-typhoidal *Salmonella enterica* isolated from normally sterile sites, 1941 to 2019 (82 articles)

**Supplemental Figure S4** - Global distribution of serogrouped isolates by serogroup by decade, global systematic review on prevalence of serogroups and serovars of non-typhoidal Salmonella enterica isolated from normally sterile sites, 1941 to 2019 (82 articles, 24,257 isolates)

**Supplemental Figure S5** - Prevalence of non-typhoidal *Salmonella enterica* serogroups isolated from normally sterile sites, by UN region, 1941-2019 (82 articles, 24,258 isolates)

**Supplemental Figure S6** - Forest plot of meta-analysis of prevalence of non-typhoidal *Salmonella enterica* from normally sterile sites of all serogrouped isolates: serogroup O:4, serogroup O:9, and other serogroups, 1941-2019 (82 articles, 24,258 isolates)

**Supplemental Figure S7** - Forest plot of meta-analysis of prevalence of non-typhoidal *Salmonella enterica* from normally sterile sites of all serogrouped isolates: serogroup O:4, serogroup O:9, and other serogroups per UN region, 1941-2019 (82 articles, 24,258 isolates)

**Supplemental Figure S8** - Forest plot of meta-analysis of prevalence of non-typhoidal *Salmonella enterica* from normally sterile sites of all serogrouped isolates: serogroup O:4, serogroup O:9, and other serogroups per age groups, 1941-2019 (82 articles, 24,258 isolates)

**Supplemental Figure S9** - Forest plot of meta-analysis of prevalence of non-typhoidal *Salmonella enterica* serovar Typhimurium, *Salmonella enterica* serovar Enteritidis and other serovars from normally sterile sites, 1941-2019 (79 articles, 23,976 isolates)

## Author contributions

Conceptualization: NH, CSM, JAC

Methodology: NH, SM, CSM, JAC

Formal analysis: NH

Data Curation: NH, CSM, JAC

Investigation: NH, SM, MB, CSM, JAC

Writing - Original Draft: NH

Writing - Review & Editing: SM, MB, CSM, JAC

Supervision: JAC

Approval of final draft: Vacc-iNTS consortium

## Research data

Data underlying this study will be made available upon publication in a repository.

