## Supplementary material for "Prevalence and distribution of non-typhoidal *Salmonella enterica* serogroups and serovars isolated from normally sterile sites: a global systematic review"

#### Supplemental 1 – Serogroup classification based on serogroups and serovars identified in the systematic review on prevalence of serogroups and serovars of non-typhoidal *Salmonella enterica* isolated from normally sterile sites, global, 1941-2019 [2]

| ***Salmonella* serogroup** | ***Salmonella* serovar** | | | | | | | | | | | | | |
|---|---|---|---|---|---|---|---|---|---|---|---|---|---|---|
| **O:2 (A)** | Group A | | | | | | | | | | | | | |
| **O:4 (B)** | Abony | Agona | Agama | Brancaster | Brandenburg | Bredeney | Chester | Coeln | Copenhagen | Derby | Haifa | Hato | Heidelberg | Java |
| | Kaapstad | Kiambu | Kisangani | Reading | Saintpaul | Sandiego | Schwarzengrund | Stanley | Stanleyville | Typhimurium | Wien | Group B | | |
| **O:7 (C1)** | Augustenborg | Bareilly | Braenderup | Choleraesuis | Colindale | Galiema | Hartford | Infantis | Irumu | Isangi | Livingstone | Mbandaka | Montevideo | Norwich |
| | Oakland | Ohio | Oranienburg | Othmarschen | Rissen | Tennessee | Thompson | Virchow | Group C | | | | | |
| **O:8 (C2-C3)** | Albany | Bardo | Blockley | Bovismorbificans | Corvallis | Hadar | Kentucky | Kottbus | Litchfield | Manhattan | Muenchen | Newport | Tshiongwe | Group C2 |
| **O:9 (D1)** | Berta | Blegdam | Dublin | Enteritidis | Javiana | Miami | Napoli | Panama | Group D | | | | | |
| **O:9,46 (D2)** | Group D2 | | | | | | | | | | | | | |
| **O:3,10 (E1)** | Amsterdam | Anatum | Give | London | Muenster | Uganda | Weltevreden | | | | | | | |
| **O:1,3,19 (E4)** | Krefeld | Senftenberg | | | | | | | | | | | | |
| **O:11 (F)** | Chandans | Rubislaw | Senegal | | | | | | | | | | | |
| **O:13 (G)** | Havana | Kedougou | Mississippi | Poona | Raus | Telelkebir | | | | | | | | |
| **O:16 (I)** | Gaminara | Hvittingfoss | | | | | | | | | | | | |
| **O:18 (K)** | Cerro | | | | | | | | | | | | | |
| **O:21 (L)** | Minnesota | | | | | | | | | | | | | |
| **O:28 (M)** | Kibusi | Telaviv | Umbilo | | | | | | | | | | | |
| **O:30 (N)** | Matopeni | Urbana | | | | | | | | | | | | |
| **O:35 (O)** | Adelaide | | | | | | | | | | | | | |
| **O:38 (P)** | Freetown | | | | | | | | | | | | | |
| **O:40 (R)** | Johannesburg | | | | | | | | | | | | | |
| **Undesignated** | Aviana | Vitiki | | | | | | | | | | | | |

#### Supplemental 2 – Bias assessment methods for the systematic review on prevalence of serogroups and serovars of non-typhoidal *Salmonella enterica* isolated from normally sterile sites, adopted from Marchello, 2022 [1]

| Grading | Definition |
|---|---|
| L | Low risk of bias |
| M | Moderate risk of bias |
| H | High risk of bias |
| U | Unknown or unavailable to assess bias |

| Domain question | Explanation | Low risk definition | High risk definition |
|---|---|---|---|
| Study design (L/H/U) | Type of study design used to ascertain NTS deaths and complications. | Active population- or household-based surveillance, prospective observational | Passive surveillance, retrospective studies, case-control, medical or laboratory records review |
| Study setting (L/H/U) | Hospital/inpatient or community/outpatient | Community or outpatient based recruitment | Admitted or inpatient hospital-based recruitment |
| Patient selection (L/H/U) | Were specific populations targeted or were all patients were eligible for recruitment | Described a systematic collection of blood culture among eligible patients | Targeted a population (e.g. children, only adults, or groups such as HIV only) or non-NTS clinical syndrome (e.g. respiratory) |
| Final year of data collection (L/M/H) | More confidence in newer studies having more robust recruitment and analytical methods | Data collection from 2010 through 2020 | Data collection 2000 and older |
| Microbiology methods (L/H/U) | Were microbiology methods fully presented, including blood culture volume and contamination. Clearly defined how infection was attributed to NTS and not *Salmonella* Typhi or Paratyphi A, B, or C. | Appropriate microbiological methods presented for ruling out typhoidal serotypes. Described blood culture volume and contamination. | Unclear or unconventional microbiological methods. No information on blood culture volumes and contamination. |

| Domain subtotal | |
|---|---|
| 0 of 3 H risk | Low risk |
| 1 of 3 H risk | Moderate risk |

| | |
|---|---|
| >1 H risk | High risk |

**Overall risk of bias**

| | |
|---|---|
| Both subtotals low risk | Low risk |
| 1 of 2 high risk | Moderate risk |
| Both subtotals high risk | High risk |

#### Supplemental 3 - Details of included articles identified in the global systematic review on prevalence of serogroups and serovars of non-typhoidal *Salmonella enterica* isolated from normally sterile sites, 1941 to 2019 (82 articles)

| First author, publication year (reference) | Country | UN region | UN sub region | Study locality | Setting | Date start data collection | Date end data collection | Methods serovar typing | Age group | Overall bias | Number of different reported serovars | Number of different serogroups | Total reported isolates |
|---|---|---|---|---|---|---|---|---|---|---|---|---|---|
| Albert, 2019 [3] | Kuwait | Asia | Western Asia | Kuwait City | Hospital-based | 01/04/2013 | 31/05/2016 | MLST | Mixed ages | H | 16 | 9 | 61 |
| Angrist, 1946 [4] | United States of America | Americas | Northern America | New York | Hospital-based | 01/01/1941 | 31/12/1944 | Agglutination test | Mixed ages | H | 3 | 3 | 11 |
| Appiah, 2021 [5] | Uganda | Africa | Eastern Africa | Apac, Tororo, Arua, Kabale, Jinja, and Mubende | Hospital-based | 01/07/2016 | 31/01/2019 | unclear | Children only (<=15y) | M | 2 | 3 | 31 |
| Asseva, 2012 [6] | Bulgaria | Europe | Eastern Europe | n/a | National surveillance | 01/01/2005 | 31/12/2010 | Agglutination test | Mixed ages | H | 6 | 5 | 33 |
| Aubry, 1992 [7] | Burundi | Africa | Eastern Africa | Bujumbura | Hospital-based | 01/01/1991 | 31/12/1991 | unclear | Adults only (>15y) | H | 2 | 3 | 54 |
| Barrios, 2017 [8] | Uruguay | Americas | South America | Montevideo | Hospital-based | 01/01/2005 | 31/12/2010 | unclear | Children only (<=15y) | H | 4 | 4 | 10 |
| Bassa, 1989 [9] | Spain | Europe | Southern Europe | Palma de Mallorca | Hospital-based | 01/01/1979 | 30/04/1988 | unclear | Mixed ages | H | 2 | 3 | 43 |
| Berkowitz, 1984 [10] | South Africa | Africa | Southern Africa | Soweto | Hospital-based | 01/01/1982 | 31/12/1982 | unclear | Children only (<=15y) | M | 10 | 5 | 47 |
| Blomberg, 2007 [11] | Tanzania | Africa | Eastern Africa | Dar es Salaam | Hospital-based | 01/08/2001 | 31/08/2002 | unclear | Children only (<=15y) | M | 3 | 4 | 37 |
| Brent, 2006 [12] | Kenya | Africa | Eastern Africa | Kilifi | Hospital-based | 01/08/1998 | 31/07/2002 | Agglutination test | Children only (<=15y) | H | 2 | 3 | 166 |
| Cheesbrough, 1997 [13] | Democratic Republic of the Congo | Africa | Middle Africa | Western | Hospital-based | 01/01/1990 | 31/12/1992 | Agglutination test | Children only (<=15y) | M | 2 | 3 | 33 |
| Chen, 1999 [14] | Taiwan | Asia | Eastern Asia | Southern | Hospital-based | 01/01/1991 | 31/12/1996 | Agglutination test | Mixed ages | H | 3 | 3 | 42 |
| Chen, 2012 [15] | Taiwan | Asia | Eastern Asia | South | Hospital-based | 01/01/1996 | 31/12/2008 | Agglutination test | Adults only (>15y) | H | 1 | 4 | 151 |
| Chu, 2014 [16] | Taiwan | Asia | Eastern Asia | Chang Gung | Hospital-based | 01/01/2004 | 31/12/2011 | unclear | Children only (<=15y) | H | 0 | 2 | 1 |
| Ciftci, 2004 [17] | Turkey | Asia | Western Asia | Ankara | Hospital-based | 01/01/1993 | 31/12/2002 | unclear | Children only (<=15y) | H | 3 | 4 | 29 |
| Cisneros-Herreros, 2005 [18] | Spain | Europe | Southern Europe | Sevilla | Hospital-based | 01/06/2001 | 17/04/2002 | unclear | Adults only (>15y) | H | 1 | 2 | 4 |
| Cisterna, 2001 [19] | Spain | Europe | Southern Europe | Bilbao | Hospital-based | 01/01/1994 | 30/09/2001 | unclear | Mixed ages | H | 1 | 2 | 82 |
| Dhanoa, 2009 [20] | Malaysia | Asia | South-eastern Asia | George Town | Hospital-based | 01/07/2002 | 31/07/2006 | Agglutination test | Mixed ages | H | 7 | 6 | 52 |
| Falay, 2016 [21] | Democratic Republic of the Congo | Africa | Middle Africa | Oriental Province | Hospital-based | 01/05/2009 | 31/05/2014 | Agglutination test | Children only (<=15y) | H | 3 | 3 | 113 |
| Feasey, 2015 [22] | Malawi | Africa | Eastern Africa | Blantyre | Hospital-based | 01/01/1998 | 31/12/2014 | Agglutination test | Mixed ages | M | 2 | 3 | 10139 |
| Galanakis, 2007 [23] | Greece | Europe | Southern Europe | Heraklion | Hospital-based | 01/01/1993 | 31/12/2002 | Agglutination test | Children only (<=15y) | H | 5 | 5 | 14 |
| Gbadoe, 2008 [24] | Togo | Africa | Western Africa | Lomé | Hospital-based | 01/01/1995 | 31/12/2004 | Agglutination test | Children only (<=15y) | H | 3 | 3 | 60 |
| Georgilis, 1997 [25] | Greece | Europe | Southern Europe | Athens | Hospital-based | 01/01/1987 | 31/12/1996 | Agglutination test | Adults only (>15y) | H | 5 | 4 | 28 |
| Gilks, 1990 [26] | Kenya | Africa | Eastern Africa | Nairobi | Hospital-based | 30/11/1988 | 15/05/1989 | unclear | Adults only (>15y) | M | 2 | 3 | 12 |
| Glaser, 1985 [27] | United States of America | Americas | Northern America | New York | Hospital-based | 01/10/1981 | 15/02/1984 | unclear | Adults only (>15y) | H | 3 | 4 | 16 |
| Gonzalez-Hevia, 1990 [28] | Spain | Europe | Southern Europe | Aviles | Laboratory-based | 01/01/1984 | 31/12/1987 | Agglutination test | Mixed ages | H | 5 | 4 | 14 |
| Gowda, 2017 [29] | Australia | Oceania | Australia and New Zealand | Queensland | Hospital-based | 01/01/2005 | 01/01/2016 | unclear | Children only (<=15y) | H | 1 | 2 | 3 |
| Grant, 1998 [30] | Côte d'Ivoire | Africa | Western Africa | Abidjan | Hospital-based | 04/12/1995 | 18/03/1996 | unclear | Adults only (>15y) | H | 1 | 2 | 11 |
| Guiraud, 2017 [31] | Burkina Faso | Africa | Western Africa | Nanoro | Hospital-based | 13/05/2013 | 12/05/2014 | Agglutination test | Children only (<=15y) | M | 4 | 4 | 67 |

| | | | | | | | | | | | | | |
|---|---|---|---|---|---|---|---|---|---|---|---|---|---|
| Gundogdu, 2017 [32] | Türkiye | Asia | Western Asia | Kayseri | Laboratory-based | 01/04/2013 | 31/01/2016 | Agglutination test | Mixed ages | M | 2 | 3 | 7 |
| Habib, 2004 [33] | Singapore | Asia | South-eastern Asia | Singapore | Hospital-based | 01/04/2001 | 31/03/2003 | unclear | Mixed ages | H | 2 | 3 | 26 |
| Harich, 2017 [34] | India | Asia | Southern Asia | Kerala | Hospital-based | 01/08/2011 | 31/07/2013 | Agglutination test | Mixed ages | H | 1 | 2 | 13 |
| Henderson, 1947 [35] | Panama | Americas | Central America | Ancon | Laboratory-based | 01/01/1942 | 31/12/1946 | Agglutination test | Mixed ages | H | 6 | 5 | 13 |
| Hsu, 2003 [36] | Taiwan | Asia | Eastern Asia | Taipei | Hospital-based | 01/09/1995 | 30/09/2001 | Agglutination test | Mixed ages | H | 0 | 5 | 93 |
| Huang, 2004 [37] | Taiwan | Asia | Eastern Asia | Kaohsiung | Hospital-based | 01/01/1996 | 31/01/2002 | Agglutination test | Children only (<=15y) | H | 2 | 3 | 45 |
| Ispahani, 2000 [38] | United Kingdom | Europe | Northern Europe | Nottingham | Hospital-based | 01/01/1980 | 31/12/1997 | Agglutination test | Mixed ages | H | 16 | 6 | 104 |
| Jones, 2008 [39] | United States of America | Americas | Northern America | 5-10 states | National surveillance | 01/01/1996 | 31/12/2006 | unclear | Mixed ages | H | 53 | 14 | 2524 |
| Kariuki, 2006 [40] | Kenya | Africa | Eastern Africa | Nairobi | Hospital-based | 01/03/2002 | 31/05/2005 | Agglutination test | Children only (<=15y) | H | 2 | 3 | 198 |
| Kassa-Kelembho, 2003 [41] | Central African Republic | Africa | Middle Africa | Bangui | Hospital-based | 01/04/1999 | 31/07/1999 | Agglutination test | Adults only (>15y) | M | 3 | 4 | 22 |
| Katiyo, 2019 [42] | United Kingdom | Europe | Northern Europe | England | National surveillance | 01/01/2004 | 31/12/2015 | Combination of methods | Mixed ages | H | 20 | 6 | 2484 |
| Keddy, 2017 [43] | South Africa | Africa | Southern Africa | Gauteng Province | Laboratory-based | 01/01/2003 | 31/12/2013 | unclear | Mixed ages | H | 4 | 4 | 4347 |
| Kedzierska, 2008 [44] | Poland | Europe | Eastern Europe | Cracow | Hospital-based | 01/01/2000 | 31/12/2006 | Agglutination test | Adults only (>15y) | H | 4 | 4 | 30 |
| Koch, 2011 [45] | Denmark | Europe | Northern Europe | North Jutland, Aarhus, Funen | Laboratory-based | 01/01/1999 | 31/12/2008 | Agglutination test | Mixed ages | M | 18 | 7 | 313 |
| Lee, 2005 [46] | Malaysia | Asia | South-eastern Asia | Kuala Lumpur | Hospital-based | 01/01/1993 | 31/12/2002 | Agglutination test | Mixed ages | H | 6 | 7 | 12 |
| Lepage, 1989 [47] | Rwanda | Africa | Eastern Africa | Kigali | Hospital-based | 15/10/1986 | 14/12/1986 | Agglutination test | Children only (<=15y) | H | 2 | 3 | 16 |
| Lepage, 1987 [48] | Rwanda | Africa | Eastern Africa | Kigali | Community-based | 16/10/1984 | 15/10/1985 | Agglutination test | Children only (<=15y) | H | 3 | 3 | 36 |
| Lester, 1991 [49] | Denmark | Europe | Northern Europe | Copenhagen | Laboratory-based | 01/01/1984 | 31/12/1988 | unclear | Mixed ages | H | 6 | 4 | 168 |
| Maltha, 2014 [50] | Burkina Faso | Africa | Western Africa | Nanoro | Hospital-based | 01/07/2012 | 31/07/2013 | unclear | Children only (<=15y) | M | 3 | 4 | 21 |
| Mandal, 1988 [51] | United Kingdom - England | Europe | Northern Europe | Manchester | Hospital-based | 01/01/1975 | 31/12/1983 | unclear | Mixed ages | H | 13 | 6 | 108 |
| Mandomando, 2015 [52] | Mozambique | Africa | Eastern Africa | Manhiça | Hospital-based | 01/01/2001 | 31/12/2014 | Combination of methods | Children only (<=15y) | M | 16 | 8 | 620 |
| Matas, 1995 [53] | Spain | Europe | Southern Europe | Barcelona | Hospital-based | 01/01/1991 | 31/12/1991 | Agglutination test | Mixed ages | H | 0 | 2 | 66 |
| Mohan, 2019 [54] | Malaysia | Asia | South-eastern Asia | Bintulu | Hospital-based | 01/01/2011 | 31/12/2016 | Agglutination test | Children only (<=15y) | H | 9 | 7 | 38 |
| Muthumbi, 2015 [55] | Kenya | Africa | Eastern Africa | Kilifi | Hospital-based | 01/08/1998 | 31/12/2014 | Agglutination test | Mixed ages | H | 2 | 3 | 351 |
| Nathoo, 1996 [56] | Zimbabwe | Africa | Eastern Africa | Harare | Hospital-based | 01/06/1993 | 31/12/1994 | Agglutination test | Children only (<=15y) | M | 1 | 2 | 10 |
| Nelson, 1982 [57] | United States of America | Americas | Northern America | St. Louis | Hospital-based | 01/01/1975 | 31/01/1981 | Agglutination test | Children only (<=15y) | H | 4 | 3 | 5 |
| Noriega, 1994 [58] | Belgium | Europe | Western Europe | Brussels | Hospital-based | 01/01/1975 | 31/12/1990 | unclear | Adults only (>15y) | H | 4 | 3 | 29 |
| Papaevangelou, 2004 [59] | Greece | Europe | Southern Europe | Athens | Hospital-based | 01/06/1990 | 31/05/2002 | Agglutination test | Children only (<=15y) | H | 5 | 5 | 119 |
| Patra, 2018 [60] | India | Asia | Southern Asia | Manipal | Hospital-based | 01/01/2012 | 31/12/2016 | Agglutination test | Mixed ages | H | 1 | 2 | 40 |
| Phoba, 2014 [61] | Democratic Republic of the Congo | Africa | Middle Africa | Bwamanda | Hospital-based | 01/11/2011 | 31/05/2012 | Agglutination test | Children only (<=15y) | M | 2 | 3 | 85 |
| Phu Huong Lan, 2016 [62] | Vietnam | Asia | South-eastern Asia | Ho Chi Minh City | Hospital-based | 01/01/2008 | 30/06/2013 | Combination of methods | Mixed ages | M | 3 | 4 | 89 |
| Phuong, 2017 [63] | Lao People's Democratic Republic | Asia | South-eastern Asia | Vientiane | Hospital-based | 01/01/2006 | 31/12/2012 | Combination of methods | Mixed ages | M | 5 | 4 | 63 |
| Preveden, 2001 [64] | Serbia | Europe | Southern Europe | Novi Sad | Hospital-based | 01/01/1991 | 31/12/1998 | unclear | Mixed ages | H | 2 | 3 | 12 |

| | | | | | | | | | | | | | |
|---|---|---|---|---|---|---|---|---|---|---|---|---|---|
| Preziosi, 2015 [65] | Mozambique | Africa | Eastern Africa | Maputo | Hospital-based | 01/09/2011 | 30/03/2014 | Agglutination test | Adults only (>15y) | H | 2 | 3 | 10 |
| Prignet, 1993 [66] | France | Europe | Southern Europe | Toulon | Hospital-based | 01/01/1972 | 31/12/1991 | unclear | Mixed ages | H | 8 | 4 | 22 |
| Ramos, 1996 [67] | Spain | Europe | Southern Europe | Madrid | Hospital-based | 01/01/1960 | 31/12/1992 | Agglutination test | Mixed ages | H | 2 | 3 | 92 |
| Raucher, 1983 [68] | United States | Americas | Northern America | New York | Hospital-based | 01/01/1981 | 31/12/1981 | Agglutination test | Children only (<=15y) | H | 5 | 3 | 7 |
| Roberts, 1993 [69] | Canada | Americas | Northern America | Vancouver | Hospital-based | 01/01/1980 | 31/01/1992 | Agglutination test | Adults only (>15y) | H | 11 | 5 | 21 |
| Secmeer, 1995 [70] | Turkey | Asia | Western Asia | Ankara | Hospital-based | 01/08/1982 | 30/09/1992 | unclear | Children only (<=15y) | H | 2 | 3 | 75 |
| Seydi, 2005 [71] | Senegal | Africa | Western Africa | Dakar | Hospital-based | 01/01/1996 | 31/12/2003 | Agglutination test | Mixed ages | H | 2 | 3 | 49 |
| Shimoni, 1999 [72] | Israel | Asia | Western Asia | Petach Tikva | Hospital-based | 01/01/1987 | 31/12/1996 | Agglutination test | Mixed ages | H | 7 | 5 | 73 |
| Sirinavin, 1999 [73] | Thailand | Asia | South-eastern Asia | Bangkok | Hospital-based | 01/01/1978 | 31/12/1994 | Agglutination test | Children only (<=15y) | H | 4 | 5 | 172 |
| Sow, 1994 [74] | Senegal | Africa | Western Africa | Dakar | Hospital-based | 01/01/1985 | 31/12/1989 | unclear | Children only (<=15y) | H | 7 | 6 | 16 |
| Still, 2020 [75] | Mali | Africa | Western Africa | Bamako | Hospital-based & Outpatient | 01/06/2002 | 31/12/2018 | Combination of methods | Children only (<=15y) | M | 3 | 3 | 682 |
| Tabu, 2012 [76] | Kenya | Africa | Eastern Africa | Asembo (rural) & Kibera (urban) | Hospital-based | 01/10/2006 | 30/09/2009 | Agglutination test | Mixed ages | H | 3 | 3 | 67 |
| Tack, 2020 [77] | Democratic Republic of the Congo | Africa | Middle Africa | Kisantu | Hospital-based | 01/01/2015 | 31/10/2017 | Agglutination test | Mixed ages | M | 5 | 6 | 896 |
| Vandenberg, 2010 [78] | Democratic Republic of the Congo | Africa | Middle Africa | Kivu Province | Hospital-based | 01/01/2002 | 31/12/2006 | Agglutination test | Children only (<=15y) | H | 4 | 4 | 191 |
| Vlieghe, 2012 [79] | Cambodia | Asia | South-eastern Asia | Phnom Penh | Hospital-based | 01/07/2007 | 30/06/2011 | Agglutination test | Mixed ages | M | 5 | 5 | 50 |
| Walsh, 2000 [80] | Malawi | Africa | Eastern Africa | Blantyre | Hospital-based | 01/09/1996 | 31/08/1997 | Agglutination test | Children only (<=15y) | M | 2 | 3 | 140 |
| Wilkens, 1997 [81] | Ghana | Africa | Western Africa | Accra | Hospital-based | 07/12/1993 | 07/03/1994 | Agglutination test | Children only (<=15y) | M | 4 | 4 | 17 |
| Yen, 2009 [82] | Taiwan | Asia | Eastern Asia | Taipei | Hospital-based | 01/01/2004 | 31/12/2006 | Agglutination test | Adults only (>15y) | H | 3 | 4 | 71 |
| Yombi, 2015 [83] | Belgium | Europe | Western Europe | Brussels | Hospital-based | 01/01/2007 | 31/12/2012 | unclear | Mixed ages | H | 2 | 3 | 20 |
| Zaidi, 1999 [84] | United States | Americas | Northern America | Boston, MA | Hospital-based | 01/01/1979 | 31/12/1995 | unclear | Children only (<=15y) | H | 5 | 4 | 138 |

#### Supplemental 4 – Descriptive characteristics by article and by isolates identified in the global systematic review on prevalence of serogroups and serovars of non-typhoidal *Salmonella enterica* isolated from normally sterile sites, 1941 to 2019

| | Included articles, n=82 | Reported isolates, n=26,280 |
|---|---|---|
| | N (%) | N (%) |
| **Data collection duration in years** | 6 (3-10) | - |
| **Number of serogroups, median (IQR)** | 3 (2-4) | - |
| **Number of serovars, median (IQR)** | 3 (2-5) | - |
| **UN regions and UN subregions** | | |
| Africa | 31 (37.8) | 18544 (70.6) |
| Eastern Africa | 15 (19.3) | 11887 (45.2) |
| Middle Africa | 6 (7.3) | 1340 (5.1) |
| Northern Africa | - | - |
| Southern Africa | 2 (2.4) | 4394 (16.7) |
| Western Africa | 8 (9.8) | 923 (3.5) |
| The Americas | 9 (10.9) | 2745 (10.4) |
| Caribbean | - | - |
| Central America | 1 (1.2) | 13 (0) |
| Northern America | 7 (8.5) | 2722 (10.4) |
| South America | 1 (1.2) | 10 (0) |
| Asia | 21 (25.6) | 1203 (4.6) |
| Central Asia | - | - |
| Eastern Asia | 6 (7.3) | 403 (1.5) |
| South-eastern Asia | 8 (9.8) | 502 (1.9) |
| Southern Asia | 2 (2.4) | 53 (0.2) |
| Western Asia | 5 (6.1) | 245 (0.9) |
| Europe | 20 (24.4) | 3785 (14.4) |
| Eastern Europe | 2 (2.4) | 63 (0.2) |
| Northern Europe | 5 (6.1) | 3177 (12.1) |
| Southern Europe | 11 (13.4) | 496 (1.9) |
| Western Europe | 2 (2.4) | 49 (0.2) |
| Oceania | 1 (1.2) | 3 (0) |
| Australia and New Zealand | 1 (1.2) | 3 (0) |
| Micronesia | - | - |
| Melanesia | - | - |
| Polynesia | - | - |
| **Setting** | | |
| Community-based | 1 (1.2) | 36 (0.1) |
| Hospital-based | 71 (86.6) | 15659 (59.6) |
| Laboratory-based | 6 (7.3) | 4862 (18.5) |
| National surveillance | 3 (3.7) | 5041 (19.2) |
| Combined | 1 (1.2) | 682 (2.6) |
| **Age group** | | |
| Adults only | 13 (15.8) | 459 (1.8) |
| Children only | 33 (40.2) | 3242 (12.3) |
| Mixed ages | 36 (43.9) | 22579 (58.9) |
| **Methods serovar typing** | | |
| Agglutination testing | 49 (59.7) | |
| Combination of methods | 5 (6.1) | |

| | |
|---|---|
| Multilocus sequence typing | 1 (1.2) |
| Unclear | 27 (32.9) |

Supplemental 5 - The proportion of non-typhoidal *Salmonella enterica* isolates by serovar for all isolates and the proportion of each serovar by serogroup identified in the global systematic review on prevalence of serogroups and serovars of non-typhoidal *Salmonella enterica* from normally sterile sites, 1941 to 2019 (26,280 isolates)

| ***Salmonella* serovar** | ***Salmonella* serogroup** | **N isolates (% of all isolates)** | **N Isolates (% in serogroup)** |
|---|---|---|---|
| Typhimurium | O:4 (B) | 14,317 (54.5) | 14,317 (93.3) |
| Enteritidis | O:9 (D1) | 6,561 (25) | 6,561 (88.8) |
| Dublin | O:9 (D1) | 524 (2) | 524 (7.1) |
| Heidelberg | O:4 (B) | 473 (1.8) | 473 (3.1) |
| Choleraesuis | O:7 (C1) | 226 (0.9) | 226 (21.2) |
| Virchow | O:7 (C1) | 218 (0.8) | 218 (20.5) |
| Isangi | O:7 (C1) | 176 (0.7) | 176 (16.5) |
| Oranienburg | O:7 (C1) | 138 (0.5) | 138 (13) |
| Newport | O:8 (C2-C3) | 121 (0.5) | 121 (48.4) |
| Poona | O:13 (G) | 95 (0.4) | 95 (91.3) |
| Montevideo | O:7 (C1) | 89 (0.3) | 89 (8.4) |
| Panama | O:9 (D1) | 82 (0.3) | 82 (1.1) |
| Infantis | O:7 (C1) | 79 (0.3) | 79 (7.4) |
| Java | O:4 (B) | 75 (0.3) | 75 (0.5) |
| Saintpaul | O:4 (B) | 71 (0.3) | 71 (0.5) |
| Javiana | O:9 (D1) | 69 (0.3) | 69 (0.9) |
| Schwarzengrund | O:4 (B) | 59 (0.2) | 59 (0.4) |
| Stanley | O:4 (B) | 58 (0.2) | 58 (0.4) |
| Serovars with <=0.1% prevalence^ | Mixed serogroups^^ | 544 (2.1) | - |
| Only reported per group | Mixed serogroups* | 286 (1.1) | - |
| Not further identified | - | 2,018 (7.7) | - |

^ List of serovars with prevalence <=0.1% is provided in Supplemental 6

^^Mixed serogroups: O:1,3,19 (E4), n=21; O:3,10 (E1), n=37; O:4 (B), n=166; O:7 (C1) n=110; O:8 (C2-C3), n=128; O:9 (D1), n=24; O:13 (G), n=9; O:30 (N), n=14; serogroups with <10 cases, n=31; undesignated, n=4

*Mixed serogroups: O:4 (B), n=129; O:7 (C1), n=28; O:8 (C2-C3), n=1; O:9 (D1), n=126; serogroups with <10 cases, n=2

#### Supplemental 6 – Serovars with prevalence ≤0.1% identified in the global systematic review on prevalence of serogroups and serovars of non-typhoidal Salmonella enterica isolated from normally sterile sites, 1941 to 2019 (26,280 isolates)

| *Salmonella* serovar | N (%) of all isolates) |
|---|---|
| Corvallis | 35 (0.1) |
| Hadar | 33 (0.1) |
| Brandenburg | 33 (0.1) |
| Sandiego | 31 (0.1) |
| Agona | 29 (0.1) |
| Bovismorbificans | 27 (0.1) |
| Braenderup | 23 (0.1) |
| Chester | 21 (0.1) |
| Berta | 17 (0.1) |
| Thompson | 17 (0.1) |
| Colindale | 16 (0.1) |
| Bareilly | 15 (0.1) |
| Krefeld | 15 (0.1) |
| Urbana | 13 (0) |
| Muenchen | 12 (0) |
| Muenster | 12 (0) |
| Ohio | 11 (0) |
| Derby | 11 (0) |
| Anatum | 11 (0) |
| Reading | 10 (0) |
| Bredeney | 9 (0) |
| Mbandaka | 8 (0) |
| Tennessee | 7 (0) |
| Rubislaw | 6 (0) |
| Senftenberg | 6 (0) |
| Uganda | 6 (0) |
| Cerro | 5 (0) |
| Copenhagen | 5 (0) |
| Kentucky | 5 (0) |
| Mississippi | 4 (0) |
| Johannesburg | 4 (0) |
| Give | 4 (0) |
| Livingstone | 4 (0) |
| Litchfield | 4 (0) |
| Hartford | 3 (0) |
| Blockley | 3 (0) |
| Gaminara | 3 (0) |
| Aviana | 3 (0) |
| Blegdam | 3 (0) |
| Haifa | 3 (0) |
| Minnesota | 3 (0) |
| Kiambu | 3 (0) |
| Adelaide | 3 (0) |
| Bardo | 2 (0) |
| Abony | 2 (0) |

| | |
|---|---|
| Albany | 2 (0) |
| Irumu | 2 (0) |
| Kedougou | 2 (0) |
| Kisangani | 2 (0) |
| Napoli | 2 (0) |
| Telelkebir | 2 (0) |
| Tshiongwe | 2 (0) |
| Weltevreden | 2 (0) |
| Miami | 2 (0) |
| Manhattan | 1 (0) |
| Agama | 1 (0) |
| Amsterdam | 1 (0) |
| Brancaster | 1 (0) |
| Chandans | 1 (0) |
| Coeln | 1 (0) |
| Freetown | 1 (0) |
| Galiema | 1 (0) |
| Hato | 1 (0) |
| Havana | 1 (0) |
| Hvittingfoss | 1 (0) |
| Kaapstad | 1 (0) |
| Kibusi | 1 (0) |
| Kottbus | 1 (0) |
| London | 1 (0) |
| Matopeni | 1 (0) |
| Oakland | 1 (0) |
| Othmarschen | 1 (0) |
| Rissen | 1 (0) |
| Senegal | 1 (0) |
| Stanleyville | 1 (0) |
| Telaviv | 1 (0) |
| Umbilo | 1 (0) |
| Vitiki | 1 (0) |
| Wien | 1 (0) |

#### Supplemental Figure 1 – PRISMA flowchart of study selection process for the global systematic review on prevalence of serogroups and serovars of non-typhoidal *Salmonella enterica* from normally sterile sites, 1941 to 2019

Legend: *Multiple reasons for exclusion possible

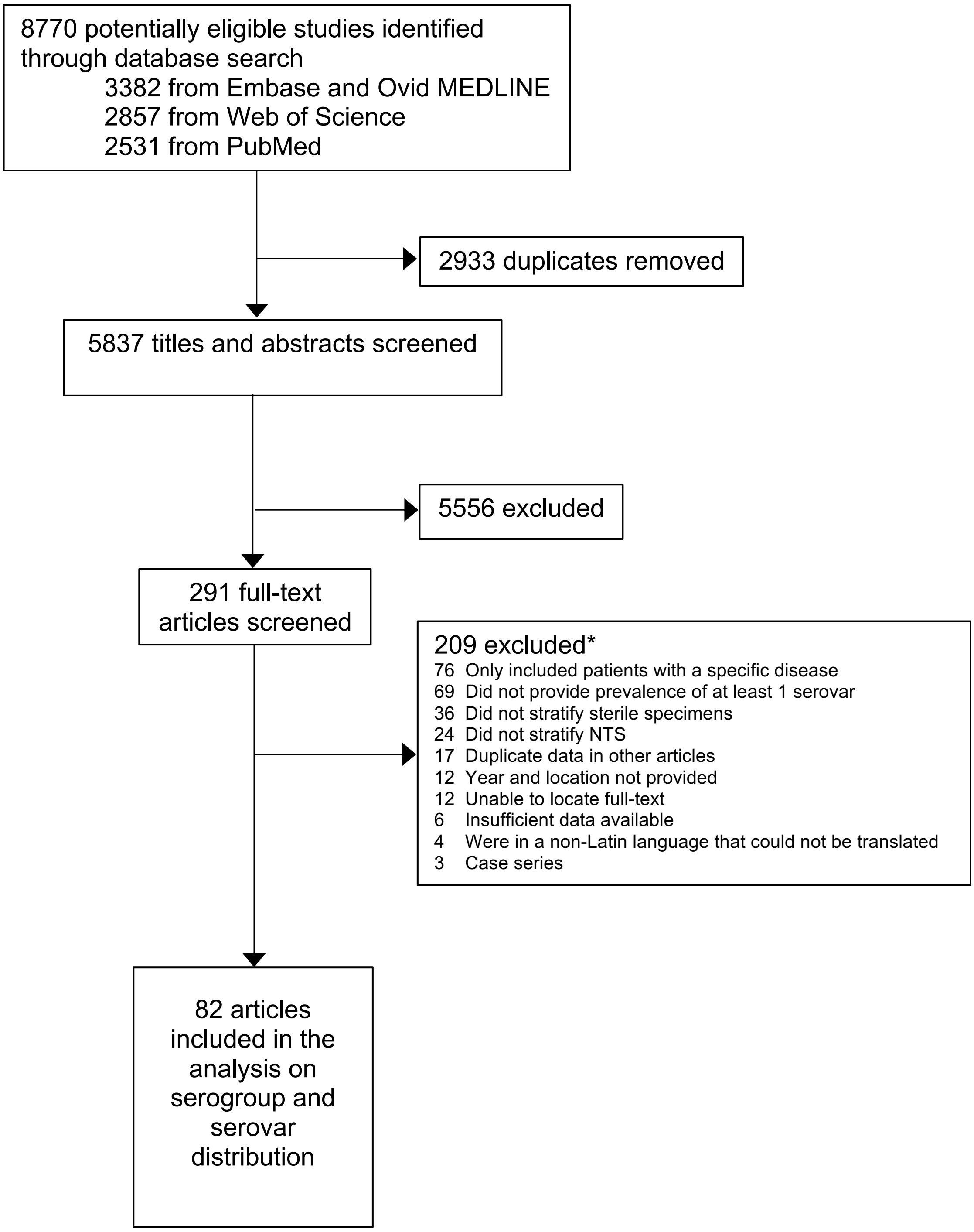

8770 potentially eligible studies identified through database search
3382 from Embase and Ovid MEDLINE
2857 from Web of Science
2531 from PubMed
2933 duplicates removed
5837 titles and abstracts screened
5556 excluded
291 full-text articles screened
209 excluded*
76 Only included patients with a specific disease
69 Did not provide prevalence of at least 1 serovar
36 Did not stratify sterile specimens
24 Did not stratify NTS
17 Duplicate data in other articles
12 Year and location not provided
12 Unable to locate full-text
6 Insufficient data available
4 Were in a non-Latin language that could not be translated
3 Case series
82 articles included in the analysis on serogroup and serovar distribution

Supplemental Figure 2 - Global distribution of number of articles per country identified in the global systematic review on prevalence of serogroups and serovars of non-typhoidal *Salmonella enterica* isolated from normally sterile sites, 1941 to 2019 (82 articles)

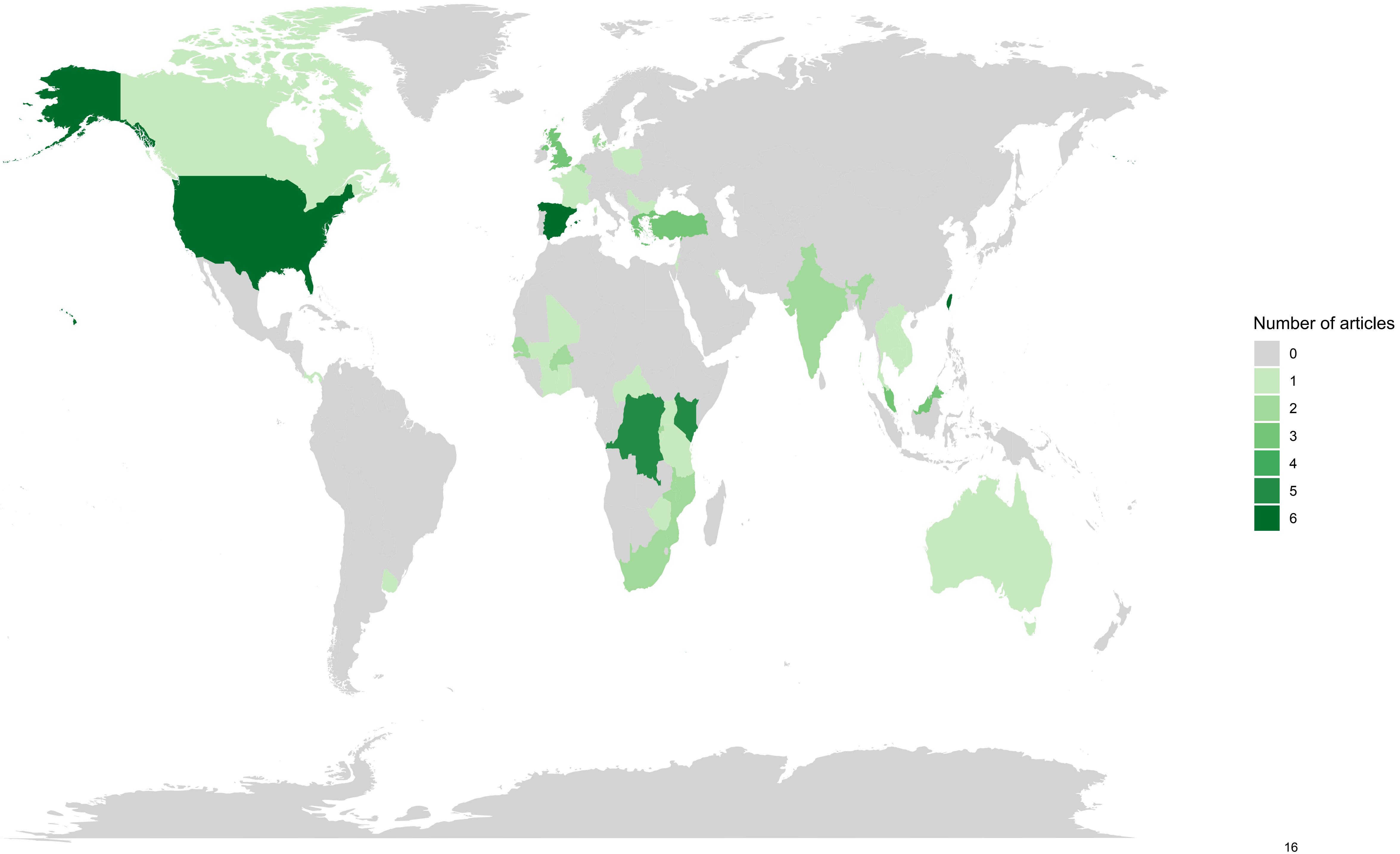
Number of articles
0
1
2
3
4
5
6

Supplemental Figure 3 – Bias assessment of the global systematic review on prevalence of serogroups and serovars of non-typhoidal *Salmonella enterica* isolated from normally sterile sites, 1941 to 2019 (82 articles)

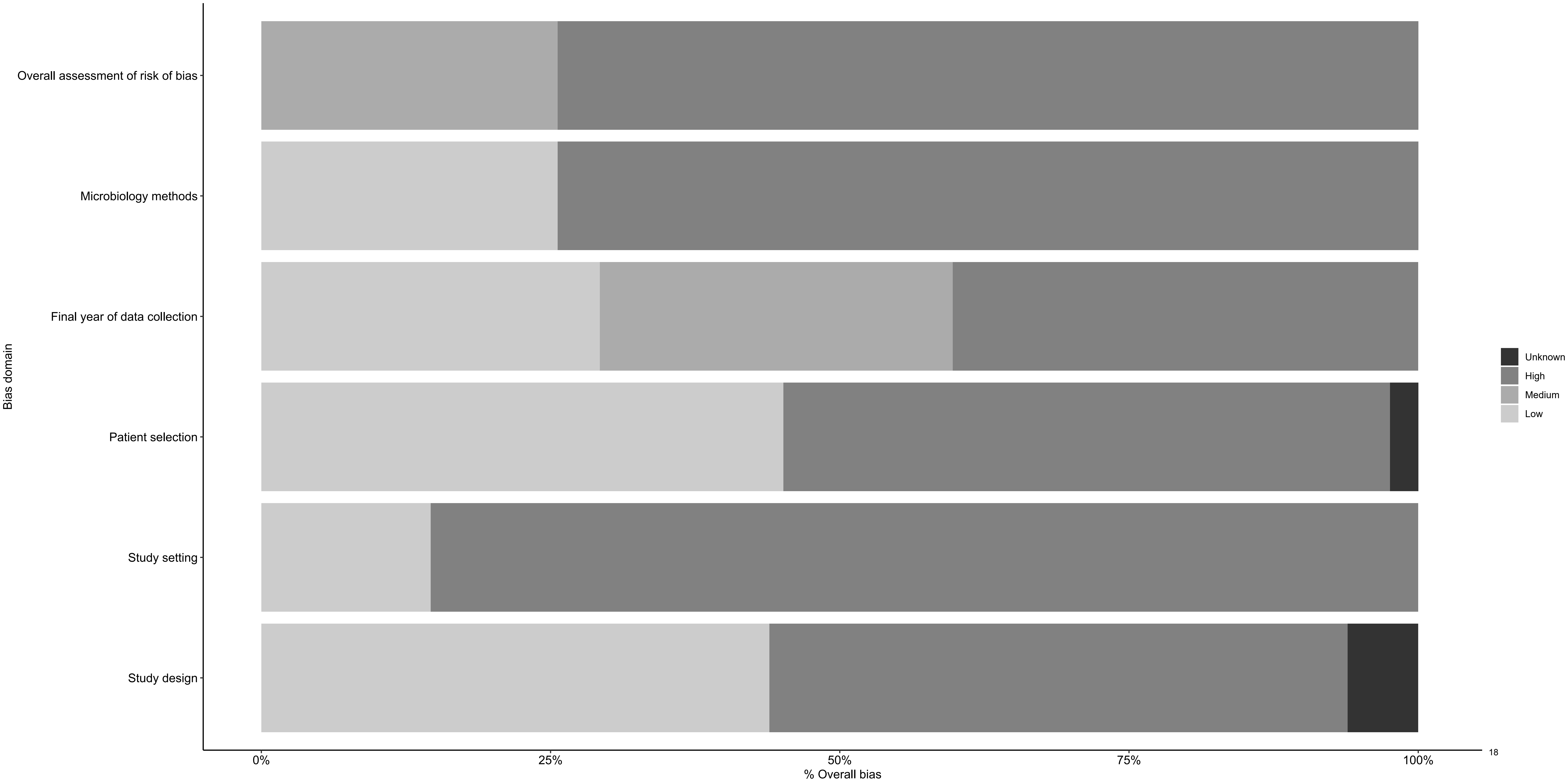
Overall assessment of risk of bias
Microbiology methods
Final year of data collection
Patient selection
Study setting
Study design
Bias domain
0%
25%
50%
75%
100%
% Overall bias
Unknown
High
Medium
Low

Supplemental Figure 4 - Global distribution of serogrouped isolates by serogroup by decade, global systematic review on prevalence of serogroups and serovars of non-typhoidal Salmonella enterica isolated from normally sterile sites, 1941 to 2019 (82 articles, 24,257 isolates)

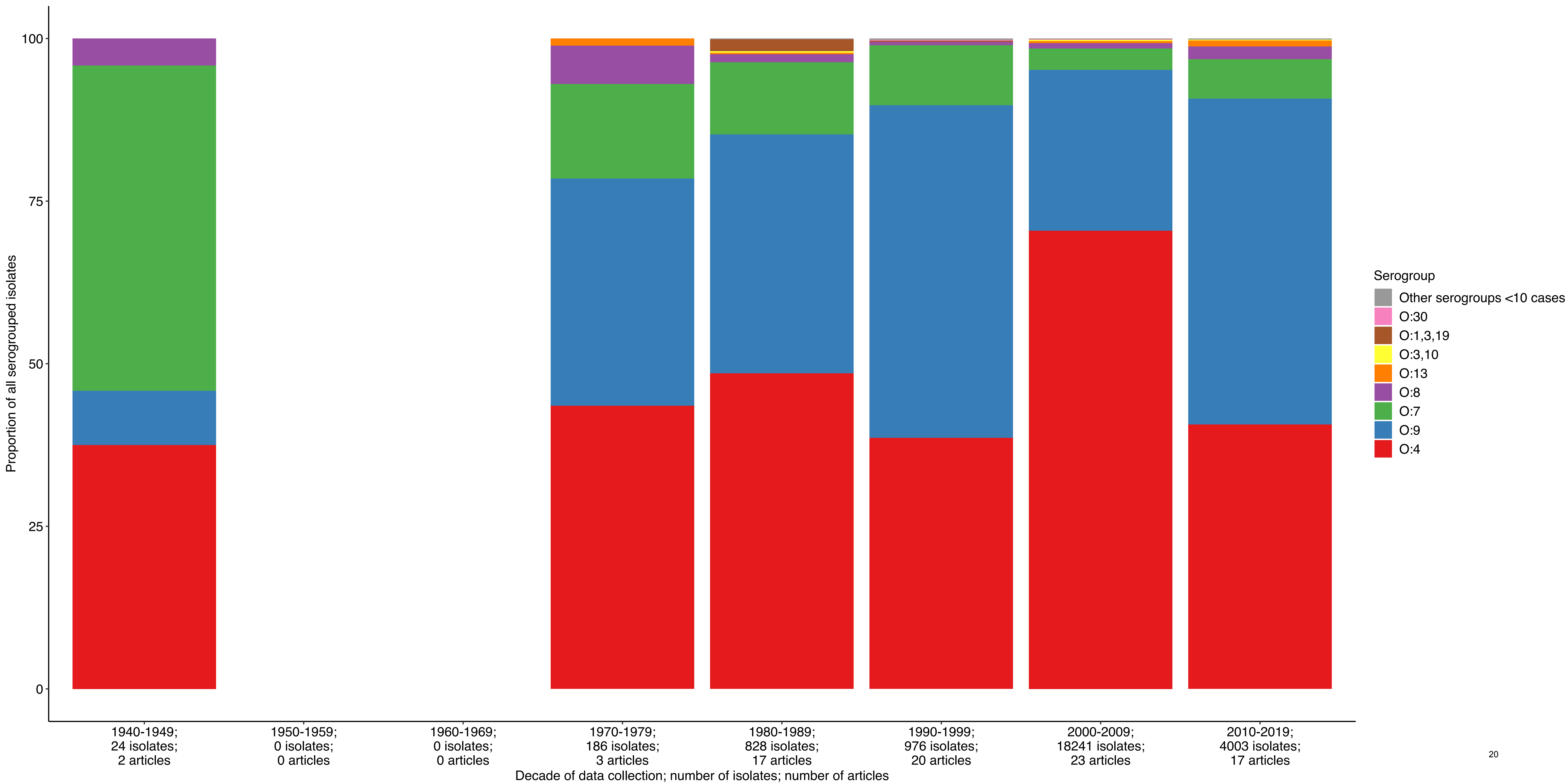
Proportion of all serogrouped isolates
100
75
50
25
0
1940-1949; 24 isolates; 2 articles
1950-1959; 0 isolates; 0 articles
1960-1969; 0 isolates; 0 articles
1970-1979; 186 isolates; 3 articles
1980-1989; 828 isolates; 17 articles
1990-1999; 976 isolates; 20 articles
2000-2009; 18241 isolates; 23 articles
2010-2019; 4003 isolates; 17 articles
Decade of data collection; number of isolates; number of articles
Serogroup
Other serogroups <10 cases
O:30
O:1,3,19
O:3,10
O:13
O:8
O:7
O:9
O:4

Supplemental Figure 5 - Prevalence of non-typhoidal *Salmonella enterica* serogroups from normally steriles, by UN region, 1941-2019 (82 articles, 24,258 isolates)

*Other serogroups with <10 cases.

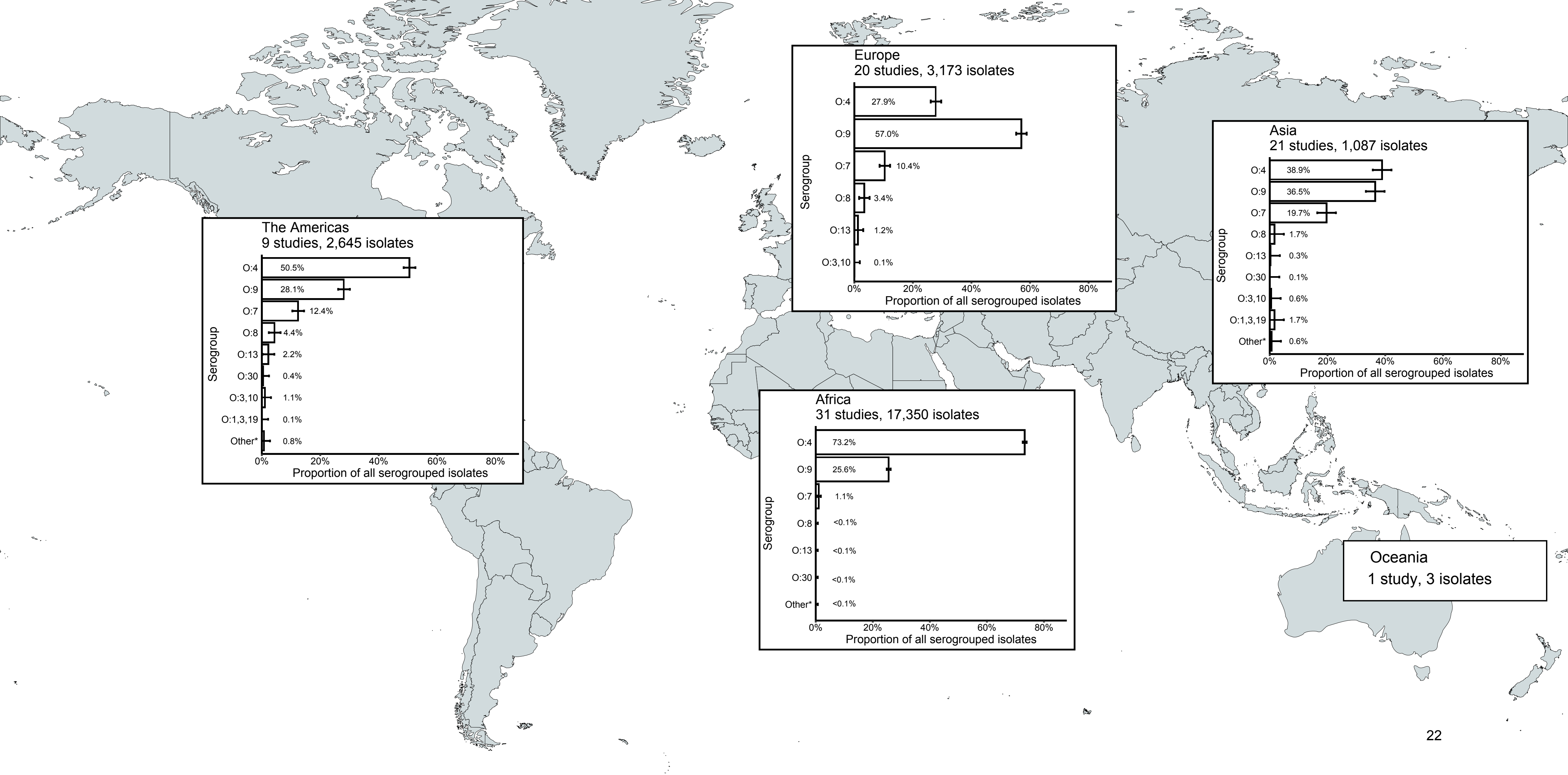
The Americas
9 studies, 2,645 isolates
O:4 50.5%
O:9 28.1%
O:7 12.4%
O:8 4.4%
O:13 2.2%
O:30 0.4%
O:3,10 1.1%
O:1,3,19 0.1%
Other* 0.8%
Serogroup
0% 20% 40% 60% 80%
Proportion of all serogrouped isolates
Europe
20 studies, 3,173 isolates
O:4 27.9%
O:9 57.0%
O:7 10.4%
O:8 3.4%
O:13 1.2%
O:3,10 0.1%
Serogroup
0% 20% 40% 60% 80%
Proportion of all serogrouped isolates
Asia
21 studies, 1,087 isolates
O:4 38.9%
O:9 36.5%
O:7 19.7%
O:8 1.7%
O:13 0.3%
O:30 0.1%
O:3,10 0.6%
O:1,3,19 1.7%
Other* 0.6%
Serogroup
0% 20% 40% 60% 80%
Proportion of all serogrouped isolates
Africa
31 studies, 17,350 isolates
O:4 73.2%
O:9 25.6%
O:7 1.1%
O:8 <0.1%
O:13 <0.1%
O:30 <0.1%
Other* <0.1%
Serogroup
0% 20% 40% 60% 80%
Proportion of all serogrouped isolates
Oceania
1 study, 3 isolates

Supplemental Figure 6 - Forest plot of meta-analysis of prevalence of non-typhoidal *Salmonella enterica* from normally sterile sites of all serogrouped isolates: serogroup O:4, serogroup O:9, and other serogroups, 1941-2019 (82 articles, 24,258 isolates)

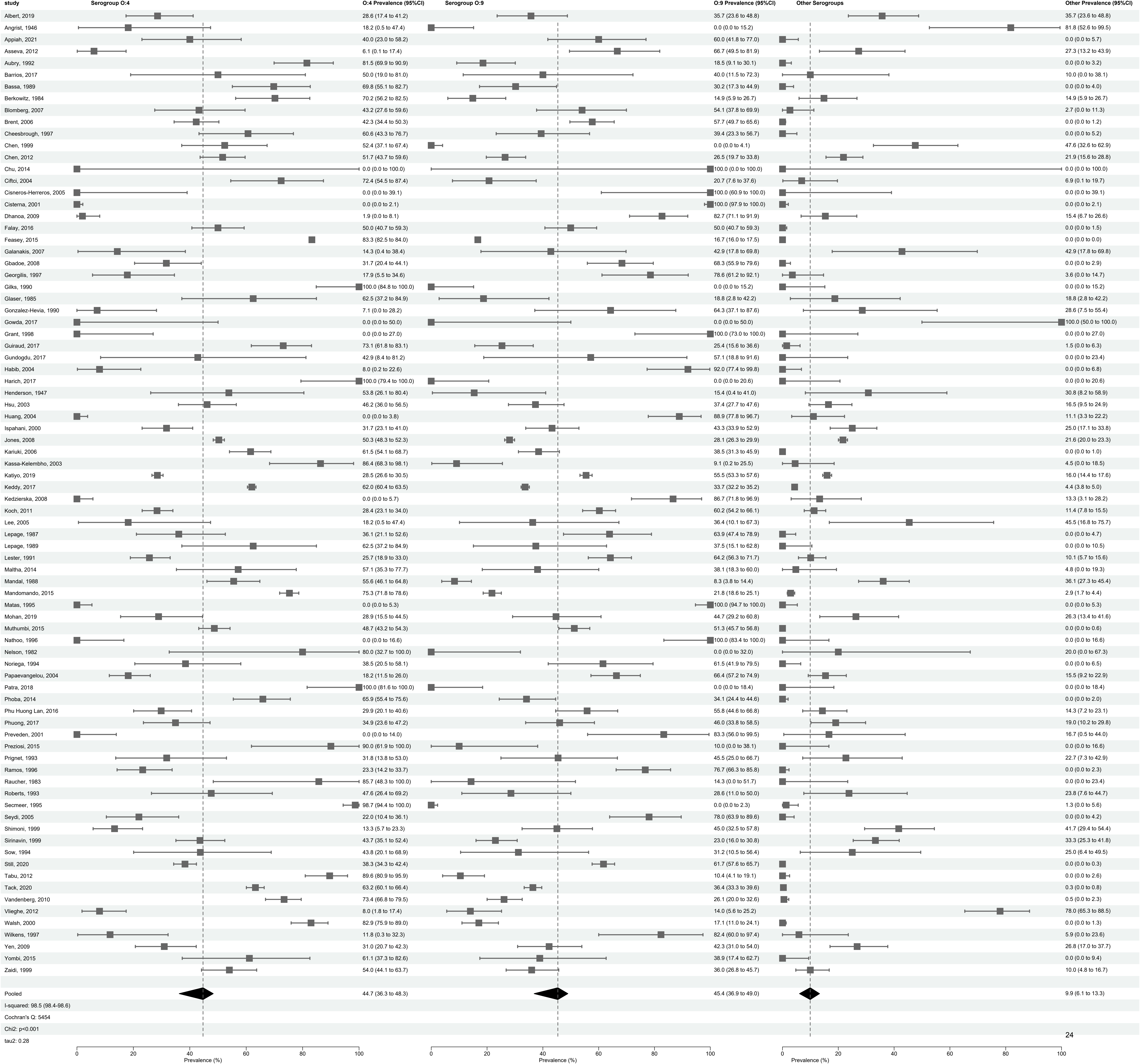
study
Serogroup O:4
O:4 Prevalence (95%CI)
Serogroup O:9
O:9 Prevalence (95%CI)
Other Serogroups
Other Prevalence (95%CI)
Albert, 2019 28.6 (17.4 to 41.2) 35.7 (23.6 to 48.8) 35.7 (23.6 to 48.8)
Angrist, 1946 18.2 (0.5 to 47.4) 0.0 (0.0 to 15.2) 81.8 (52.6 to 99.5)
Appiah, 2021 40.0 (23.0 to 58.2) 60.0 (41.8 to 77.0) 0.0 (0.0 to 5.7)
Asseva, 2012 6.1 (0.1 to 17.4) 66.7 (49.5 to 81.9) 27.3 (13.2 to 43.9)
Aubry, 1992 81.5 (69.9 to 90.9) 18.5 (9.1 to 30.1) 0.0 (0.0 to 3.2)
Barrios, 2017 50.0 (19.0 to 81.0) 40.0 (11.5 to 72.3) 10.0 (0.0 to 38.1)
Bassa, 1989 69.8 (55.1 to 82.7) 30.2 (17.3 to 44.9) 0.0 (0.0 to 4.0)
Berkowitz, 1984 70.2 (56.2 to 82.5) 14.9 (5.9 to 26.7) 14.9 (5.9 to 26.7)
Blomberg, 2007 43.2 (27.6 to 59.6) 54.1 (37.8 to 69.9) 2.7 (0.0 to 11.3)
Brent, 2006 42.3 (34.4 to 50.3) 57.7 (49.7 to 65.6) 0.0 (0.0 to 1.2)
Cheesbrough, 1997 60.6 (43.3 to 76.7) 39.4 (23.3 to 56.7) 0.0 (0.0 to 5.2)
Chen, 1999 52.4 (37.1 to 67.4) 0.0 (0.0 to 4.1) 47.6 (32.6 to 62.9)
Chen, 2012 51.7 (43.7 to 59.6) 26.5 (19.7 to 33.8) 21.9 (15.6 to 28.8)
Chu, 2014 0.0 (0.0 to 100.0) 100.0 (0.0 to 100.0) 0.0 (0.0 to 100.0)
Ciftci, 2004 72.4 (54.5 to 87.4) 20.7 (7.6 to 37.6) 6.9 (0.1 to 19.7)
Cisneros-Herreros, 2005 0.0 (0.0 to 39.1) 100.0 (60.9 to 100.0) 0.0 (0.0 to 39.1)
Cisterna, 2001 0.0 (0.0 to 2.1) 100.0 (97.9 to 100.0) 0.0 (0.0 to 2.1)
Dhanoa, 2009 1.9 (0.0 to 8.1) 82.7 (71.1 to 91.9) 15.4 (6.7 to 26.6)
Falay, 2016 50.0 (40.7 to 59.3) 50.0 (40.7 to 59.3) 0.0 (0.0 to 1.5)
Feasey, 2015 83.3 (82.5 to 84.0) 16.7 (16.0 to 17.5) 0.0 (0.0 to 0.0)
Galanakis, 2007 14.3 (0.4 to 38.4) 42.9 (17.8 to 69.8) 42.9 (17.8 to 69.8)
Gbadoe, 2008 31.7 (20.4 to 44.1) 68.3 (55.9 to 79.6) 0.0 (0.0 to 2.9)
Georgilis, 1997 17.9 (5.5 to 34.6) 78.6 (61.2 to 92.1) 3.6 (0.0 to 14.7)
Gilks, 1990 100.0 (84.8 to 100.0) 0.0 (0.0 to 15.2) 0.0 (0.0 to 15.2)
Glaser, 1985 62.5 (37.2 to 84.9) 18.8 (2.8 to 42.2) 18.8 (2.8 to 42.2)
Gonzalez-Hevia, 1990 7.1 (0.0 to 28.2) 64.3 (37.1 to 87.6) 28.6 (7.5 to 55.4)
Gowda, 2017 0.0 (0.0 to 50.0) 0.0 (0.0 to 50.0) 100.0 (50.0 to 100.0)
Grant, 1998 0.0 (0.0 to 27.0) 100.0 (73.0 to 100.0) 0.0 (0.0 to 27.0)
Guiraud, 2017 73.1 (61.8 to 83.1) 25.4 (15.6 to 36.6) 1.5 (0.0 to 6.3)
Gundogdu, 2017 42.9 (8.4 to 81.2) 57.1 (18.8 to 91.6) 0.0 (0.0 to 23.4)
Habib, 2004 8.0 (0.2 to 22.6) 92.0 (77.4 to 99.8) 0.0 (0.0 to 6.8)
Harich, 2017 100.0 (79.4 to 100.0) 0.0 (0.0 to 20.6) 0.0 (0.0 to 20.6)
Henderson, 1947 53.8 (26.1 to 80.4) 15.4 (0.4 to 41.0) 30.8 (8.2 to 58.9)
Hsu, 2003 46.2 (36.0 to 56.5) 37.4 (27.7 to 47.6) 16.5 (9.5 to 24.9)
Huang, 2004 0.0 (0.0 to 3.8) 88.9 (77.8 to 96.7) 11.1 (3.3 to 22.2)
Ispahani, 2000 31.7 (23.1 to 41.0) 43.3 (33.9 to 52.9) 25.0 (17.1 to 33.8)
Jones, 2008 50.3 (48.3 to 52.3) 28.1 (26.3 to 29.9) 21.6 (20.0 to 23.3)
Kariuki, 2006 61.5 (54.1 to 68.7) 38.5 (31.3 to 45.9) 0.0 (0.0 to 1.0)
Kassa-Kelembho, 2003 86.4 (68.3 to 98.1) 9.1 (0.2 to 25.5) 4.5 (0.0 to 18.5)
Katiyo, 2019 28.5 (26.6 to 30.5) 55.5 (53.3 to 57.6) 16.0 (14.4 to 17.6)
Keddy, 2017 62.0 (60.4 to 63.5) 33.7 (32.2 to 35.2) 4.4 (3.8 to 5.0)
Kedzierska, 2008 0.0 (0.0 to 5.7) 86.7 (71.8 to 96.9) 13.3 (3.1 to 28.2)
Koch, 2011 28.4 (23.1 to 34.0) 60.2 (54.2 to 66.1) 11.4 (7.8 to 15.5)
Lee, 2005 18.2 (0.5 to 47.4) 36.4 (10.1 to 67.3) 45.5 (16.8 to 75.7)
Lepage, 1987 36.1 (21.1 to 52.6) 63.9 (47.4 to 78.9) 0.0 (0.0 to 4.7)
Lepage, 1989 62.5 (37.2 to 84.9) 37.5 (15.1 to 62.8) 0.0 (0.0 to 10.5)
Lester, 1991 25.7 (18.9 to 33.0) 64.2 (56.3 to 71.7) 10.1 (5.7 to 15.6)
Maltha, 2014 57.1 (35.3 to 77.7) 38.1 (18.3 to 60.0) 4.8 (0.0 to 19.3)
Mandal, 1988 55.6 (46.1 to 64.8) 8.3 (3.8 to 14.4) 36.1 (27.3 to 45.4)
Mandomando, 2015 75.3 (71.8 to 78.6) 21.8 (18.6 to 25.1) 2.9 (1.7 to 4.4)
Matas, 1995 0.0 (0.0 to 5.3) 100.0 (94.7 to 100.0) 0.0 (0.0 to 5.3)
Mohan, 2019 28.9 (15.5 to 44.5) 44.7 (29.2 to 60.8) 26.3 (13.4 to 41.6)
Muthumbi, 2015 48.7 (43.2 to 54.3) 51.3 (45.7 to 56.8) 0.0 (0.0 to 0.6)
Nathoo, 1996 0.0 (0.0 to 16.6) 100.0 (83.4 to 100.0) 0.0 (0.0 to 16.6)
Nelson, 1982 80.0 (32.7 to 100.0) 0.0 (0.0 to 32.0) 20.0 (0.0 to 67.3)
Noriega, 1994 38.5 (20.5 to 58.1) 61.5 (41.9 to 79.5) 0.0 (0.0 to 6.5)
Papaevangelou, 2004 18.2 (11.5 to 26.0) 66.4 (57.2 to 74.9) 15.5 (9.2 to 22.9)
Patra, 2018 100.0 (81.6 to 100.0) 0.0 (0.0 to 18.4) 0.0 (0.0 to 18.4)
Phoba, 2014 65.9 (55.4 to 75.6) 34.1 (24.4 to 44.6) 0.0 (0.0 to 2.0)
Phu Huong Lan, 2016 29.9 (20.1 to 40.6) 55.8 (44.6 to 66.8) 14.3 (7.2 to 23.1)
Phuong, 2017 34.9 (23.6 to 47.2) 46.0 (33.8 to 58.5) 19.0 (10.2 to 29.8)
Preveden, 2001 0.0 (0.0 to 14.0) 83.3 (56.0 to 99.5) 16.7 (0.5 to 44.0)
Preziosi, 2015 90.0 (61.9 to 100.0) 10.0 (0.0 to 38.1) 0.0 (0.0 to 16.6)
Prignet, 1993 31.8 (13.8 to 53.0) 45.5 (25.0 to 66.7) 22.7 (7.3 to 42.9)
Ramos, 1996 23.3 (14.2 to 33.7) 76.7 (66.3 to 85.8) 0.0 (0.0 to 2.3)
Raucher, 1983 85.7 (48.3 to 100.0) 14.3 (0.0 to 51.7) 0.0 (0.0 to 23.4)
Roberts, 1993 47.6 (26.4 to 69.2) 28.6 (11.0 to 50.0) 23.8 (7.6 to 44.7)
Secmeer, 1995 98.7 (94.4 to 100.0) 0.0 (0.0 to 2.3) 1.3 (0.0 to 5.6)
Seydi, 2005 22.0 (10.4 to 36.1) 78.0 (63.9 to 89.6) 0.0 (0.0 to 4.2)
Shimoni, 1999 13.3 (5.7 to 23.3) 45.0 (32.5 to 57.8) 41.7 (29.4 to 54.4)
Sirinavin, 1999 43.7 (35.1 to 52.4) 23.0 (16.0 to 30.8) 33.3 (25.3 to 41.8)
Sow, 1994 43.8 (20.1 to 68.9) 31.2 (10.5 to 56.4) 25.0 (6.4 to 49.5)
Still, 2020 38.3 (34.3 to 42.4) 61.7 (57.6 to 65.7) 0.0 (0.0 to 0.3)
Tabu, 2012 89.6 (80.9 to 95.9) 10.4 (4.1 to 19.1) 0.0 (0.0 to 2.6)
Tack, 2020 63.2 (60.1 to 66.4) 36.4 (33.3 to 39.6) 0.3 (0.0 to 0.8)
Vandenberg, 2010 73.4 (66.8 to 79.5) 26.1 (20.0 to 32.6) 0.5 (0.0 to 2.3)
Vlieghe, 2012 8.0 (1.8 to 17.4) 14.0 (5.6 to 25.2) 78.0 (65.3 to 88.5)
Walsh, 2000 82.9 (75.9 to 89.0) 17.1 (11.0 to 24.1) 0.0 (0.0 to 1.3)
Wilkens, 1997 11.8 (0.3 to 32.3) 82.4 (60.0 to 97.4) 5.9 (0.0 to 23.6)
Yen, 2009 31.0 (20.7 to 42.3) 42.3 (31.0 to 54.0) 26.8 (17.0 to 37.7)
Yombi, 2015 61.1 (37.3 to 82.6) 38.9 (17.4 to 62.7) 0.0 (0.0 to 9.4)
Zaidi, 1999 54.0 (44.1 to 63.7) 36.0 (26.8 to 45.7) 10.0 (4.8 to 16.7)
Pooled 44.7 (36.3 to 48.3) 45.4 (36.9 to 49.0) 9.9 (6.1 to 13.3)
I-squared: 98.5 (98.4-98.6)
Cochran's Q: 5454
Chi2: p<0.001
tau2: 0.28
0 20 40 60 80 100
Prevalence (%)
0 20 40 60 80 100
Prevalence (%)
0 20 40 60 80 100
Prevalence (%)

Supplemental Figure 7 - Forest plot of meta-analysis of prevalence of non-typhoidal *Salmonella enterica* from normally sterile sites of all serogrouped isolates: serogroup O:4, serogroup O:9, and other serogroups per UN region, 1941-2019 (82 articles, 24,258 isolates)

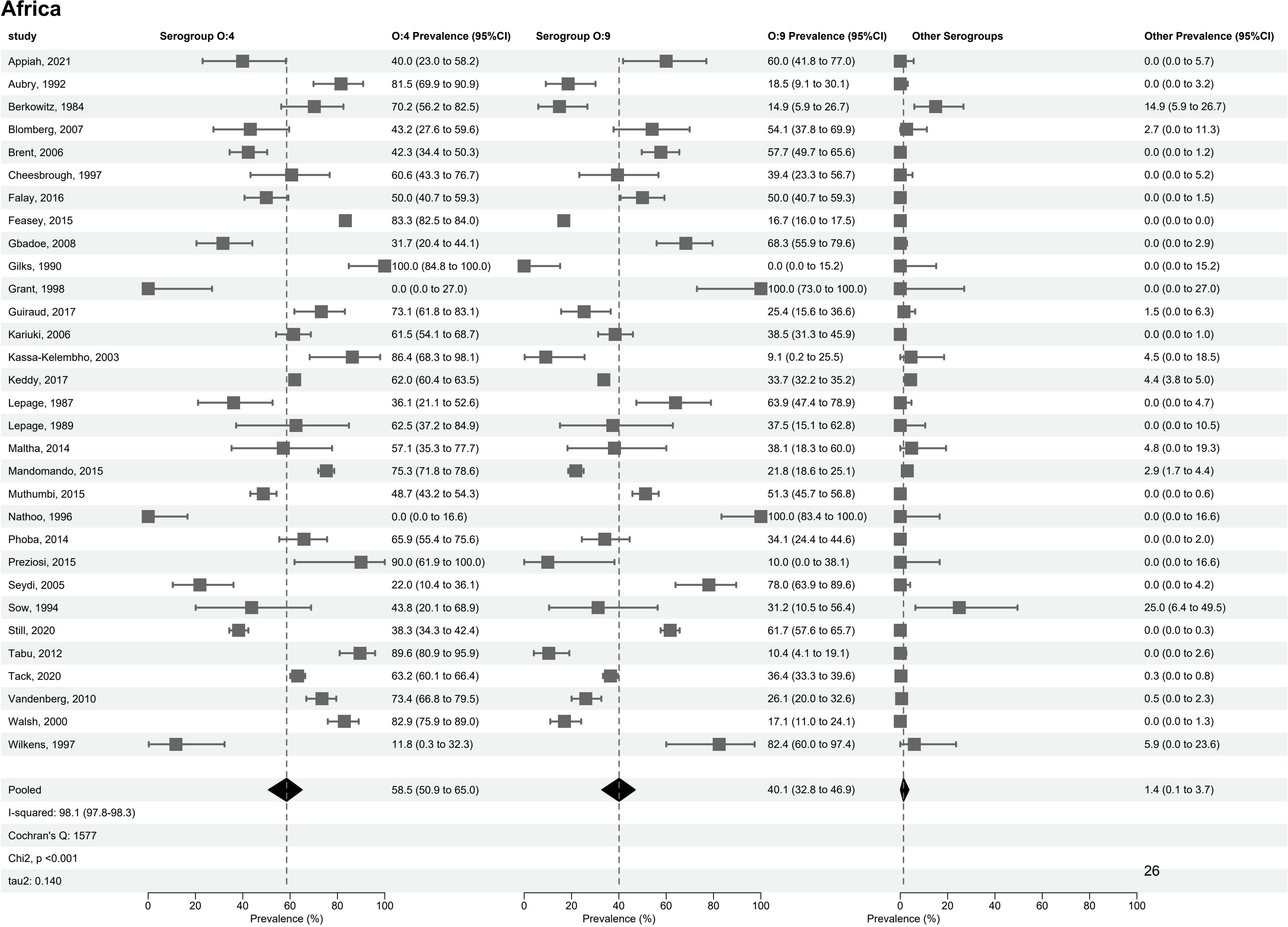
Africa
study
Serogroup O:4
O:4 Prevalence (95%CI)
Serogroup O:9
O:9 Prevalence (95%CI)
Other Serogroups
Other Prevalence (95%CI)
Appiah, 2021 40.0 (23.0 to 58.2) 60.0 (41.8 to 77.0) 0.0 (0.0 to 5.7)
Aubry, 1992 81.5 (69.9 to 90.9) 18.5 (9.1 to 30.1) 0.0 (0.0 to 3.2)
Berkowitz, 1984 70.2 (56.2 to 82.5) 14.9 (5.9 to 26.7) 14.9 (5.9 to 26.7)
Blomberg, 2007 43.2 (27.6 to 59.6) 54.1 (37.8 to 69.9) 2.7 (0.0 to 11.3)
Brent, 2006 42.3 (34.4 to 50.3) 57.7 (49.7 to 65.6) 0.0 (0.0 to 1.2)
Cheesbrough, 1997 60.6 (43.3 to 76.7) 39.4 (23.3 to 56.7) 0.0 (0.0 to 5.2)
Falay, 2016 50.0 (40.7 to 59.3) 50.0 (40.7 to 59.3) 0.0 (0.0 to 1.5)
Feasey, 2015 83.3 (82.5 to 84.0) 16.7 (16.0 to 17.5) 0.0 (0.0 to 0.0)
Gbadoe, 2008 31.7 (20.4 to 44.1) 68.3 (55.9 to 79.6) 0.0 (0.0 to 2.9)
Gilks, 1990 100.0 (84.8 to 100.0) 0.0 (0.0 to 15.2) 0.0 (0.0 to 15.2)
Grant, 1998 0.0 (0.0 to 27.0) 100.0 (73.0 to 100.0) 0.0 (0.0 to 27.0)
Guiraud, 2017 73.1 (61.8 to 83.1) 25.4 (15.6 to 36.6) 1.5 (0.0 to 6.3)
Kariuki, 2006 61.5 (54.1 to 68.7) 38.5 (31.3 to 45.9) 0.0 (0.0 to 1.0)
Kassa-Kelembho, 2003 86.4 (68.3 to 98.1) 9.1 (0.2 to 25.5) 4.5 (0.0 to 18.5)
Keddy, 2017 62.0 (60.4 to 63.5) 33.7 (32.2 to 35.2) 4.4 (3.8 to 5.0)
Lepage, 1987 36.1 (21.1 to 52.6) 63.9 (47.4 to 78.9) 0.0 (0.0 to 4.7)
Lepage, 1989 62.5 (37.2 to 84.9) 37.5 (15.1 to 62.8) 0.0 (0.0 to 10.5)
Maltha, 2014 57.1 (35.3 to 77.7) 38.1 (18.3 to 60.0) 4.8 (0.0 to 19.3)
Mandomando, 2015 75.3 (71.8 to 78.6) 21.8 (18.6 to 25.1) 2.9 (1.7 to 4.4)
Muthumbi, 2015 48.7 (43.2 to 54.3) 51.3 (45.7 to 56.8) 0.0 (0.0 to 0.6)
Nathoo, 1996 0.0 (0.0 to 16.6) 100.0 (83.4 to 100.0) 0.0 (0.0 to 16.6)
Phoba, 2014 65.9 (55.4 to 75.6) 34.1 (24.4 to 44.6) 0.0 (0.0 to 2.0)
Preziosi, 2015 90.0 (61.9 to 100.0) 10.0 (0.0 to 38.1) 0.0 (0.0 to 16.6)
Seydi, 2005 22.0 (10.4 to 36.1) 78.0 (63.9 to 89.6) 0.0 (0.0 to 4.2)
Sow, 1994 43.8 (20.1 to 68.9) 31.2 (10.5 to 56.4) 25.0 (6.4 to 49.5)
Still, 2020 38.3 (34.3 to 42.4) 61.7 (57.6 to 65.7) 0.0 (0.0 to 0.3)
Tabu, 2012 89.6 (80.9 to 95.9) 10.4 (4.1 to 19.1) 0.0 (0.0 to 2.6)
Tack, 2020 63.2 (60.1 to 66.4) 36.4 (33.3 to 39.6) 0.3 (0.0 to 0.8)
Vandenberg, 2010 73.4 (66.8 to 79.5) 26.1 (20.0 to 32.6) 0.5 (0.0 to 2.3)
Walsh, 2000 82.9 (75.9 to 89.0) 17.1 (11.0 to 24.1) 0.0 (0.0 to 1.3)
Wilkens, 1997 11.8 (0.3 to 32.3) 82.4 (60.0 to 97.4) 5.9 (0.0 to 23.6)
Pooled 58.5 (50.9 to 65.0) 40.1 (32.8 to 46.9) 1.4 (0.1 to 3.7)
I-squared: 98.1 (97.8-98.3)
Cochran's Q: 1577
Chi2, p <0.001
tau2: 0.140
0 20 40 60 80 100
Prevalence (%)
0 20 40 60 80 100
Prevalence (%)
0 20 40 60 80 100
Prevalence (%)

### The Americas

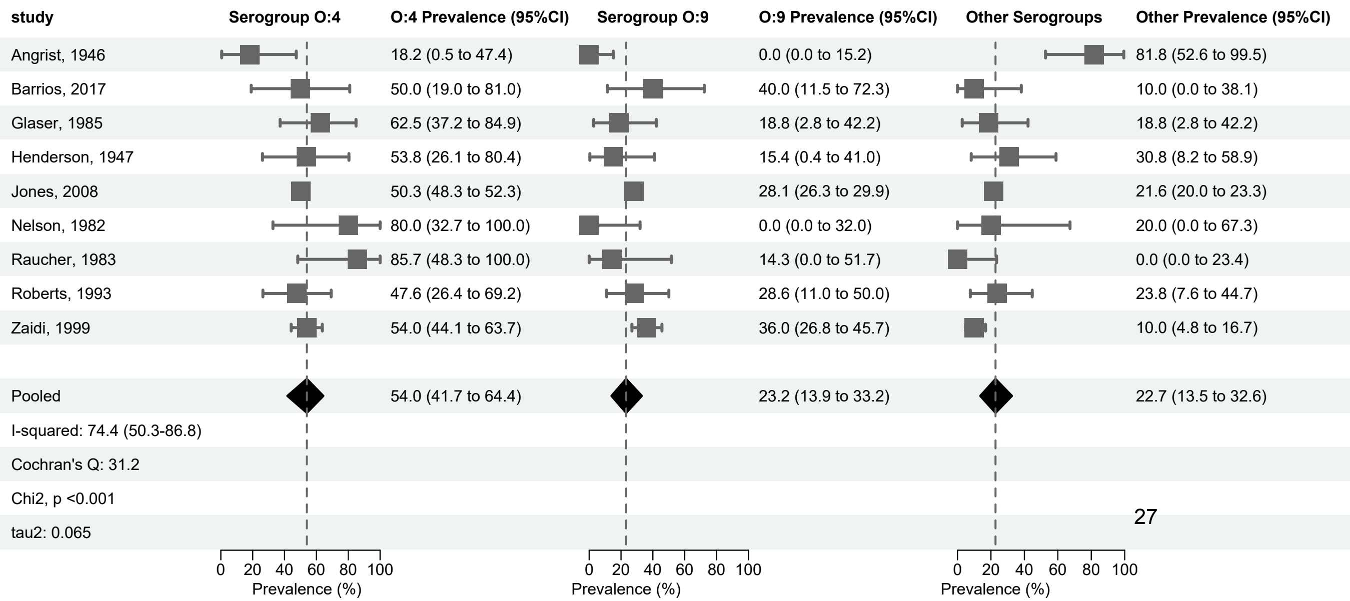

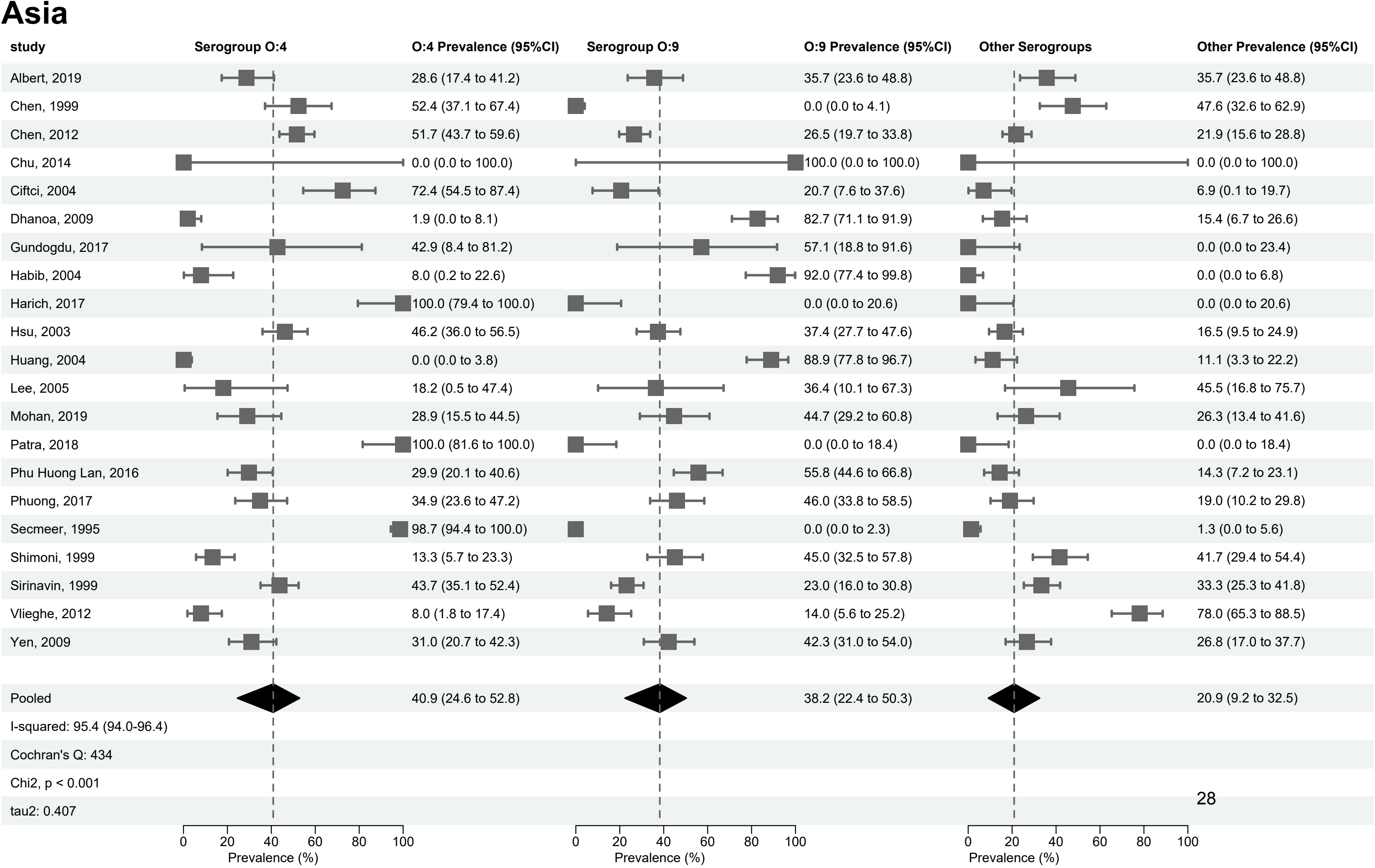
Asia
study
Serogroup O:4
O:4 Prevalence (95%CI)
Serogroup O:9
O:9 Prevalence (95%CI)
Other Serogroups
Other Prevalence (95%CI)
Albert, 2019 28.6 (17.4 to 41.2) 35.7 (23.6 to 48.8) 35.7 (23.6 to 48.8)
Chen, 1999 52.4 (37.1 to 67.4) 0.0 (0.0 to 4.1) 47.6 (32.6 to 62.9)
Chen, 2012 51.7 (43.7 to 59.6) 26.5 (19.7 to 33.8) 21.9 (15.6 to 28.8)
Chu, 2014 0.0 (0.0 to 100.0) 100.0 (0.0 to 100.0) 0.0 (0.0 to 100.0)
Ciftci, 2004 72.4 (54.5 to 87.4) 20.7 (7.6 to 37.6) 6.9 (0.1 to 19.7)
Dhanoa, 2009 1.9 (0.0 to 8.1) 82.7 (71.1 to 91.9) 15.4 (6.7 to 26.6)
Gundogdu, 2017 42.9 (8.4 to 81.2) 57.1 (18.8 to 91.6) 0.0 (0.0 to 23.4)
Habib, 2004 8.0 (0.2 to 22.6) 92.0 (77.4 to 99.8) 0.0 (0.0 to 6.8)
Harich, 2017 100.0 (79.4 to 100.0) 0.0 (0.0 to 20.6) 0.0 (0.0 to 20.6)
Hsu, 2003 46.2 (36.0 to 56.5) 37.4 (27.7 to 47.6) 16.5 (9.5 to 24.9)
Huang, 2004 0.0 (0.0 to 3.8) 88.9 (77.8 to 96.7) 11.1 (3.3 to 22.2)
Lee, 2005 18.2 (0.5 to 47.4) 36.4 (10.1 to 67.3) 45.5 (16.8 to 75.7)
Mohan, 2019 28.9 (15.5 to 44.5) 44.7 (29.2 to 60.8) 26.3 (13.4 to 41.6)
Patra, 2018 100.0 (81.6 to 100.0) 0.0 (0.0 to 18.4) 0.0 (0.0 to 18.4)
Phu Huong Lan, 2016 29.9 (20.1 to 40.6) 55.8 (44.6 to 66.8) 14.3 (7.2 to 23.1)
Phuong, 2017 34.9 (23.6 to 47.2) 46.0 (33.8 to 58.5) 19.0 (10.2 to 29.8)
Secmeer, 1995 98.7 (94.4 to 100.0) 0.0 (0.0 to 2.3) 1.3 (0.0 to 5.6)
Shimoni, 1999 13.3 (5.7 to 23.3) 45.0 (32.5 to 57.8) 41.7 (29.4 to 54.4)
Sirinavin, 1999 43.7 (35.1 to 52.4) 23.0 (16.0 to 30.8) 33.3 (25.3 to 41.8)
Vlieghe, 2012 8.0 (1.8 to 17.4) 14.0 (5.6 to 25.2) 78.0 (65.3 to 88.5)
Yen, 2009 31.0 (20.7 to 42.3) 42.3 (31.0 to 54.0) 26.8 (17.0 to 37.7)
Pooled 40.9 (24.6 to 52.8) 38.2 (22.4 to 50.3) 20.9 (9.2 to 32.5)
I-squared: 95.4 (94.0-96.4)
Cochran's Q: 434
Chi2, p < 0.001
tau2: 0.407
0 20 40 60 80 100
Prevalence (%)
0 20 40 60 80 100
Prevalence (%)
0 20 40 60 80 100
Prevalence (%)

### Europe

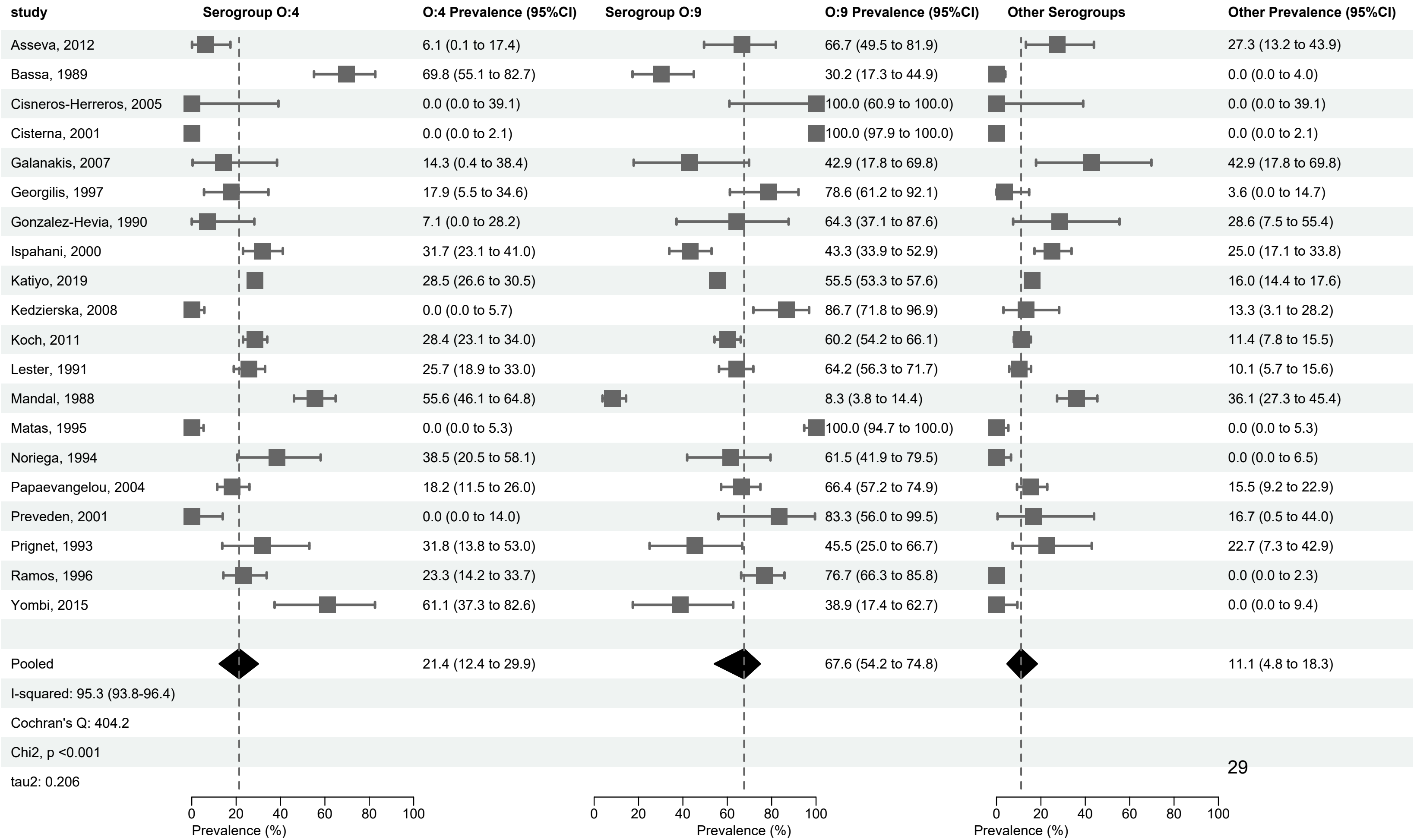
study
Serogroup O:4
O:4 Prevalence (95%CI)
Serogroup O:9
O:9 Prevalence (95%CI)
Other Serogroups
Other Prevalence (95%CI)
Asseva, 2012 6.1 (0.1 to 17.4) 66.7 (49.5 to 81.9) 27.3 (13.2 to 43.9)
Bassa, 1989 69.8 (55.1 to 82.7) 30.2 (17.3 to 44.9) 0.0 (0.0 to 4.0)
Cisneros-Herreros, 2005 0.0 (0.0 to 39.1) 100.0 (60.9 to 100.0) 0.0 (0.0 to 39.1)
Cisterna, 2001 0.0 (0.0 to 2.1) 100.0 (97.9 to 100.0) 0.0 (0.0 to 2.1)
Galanakis, 2007 14.3 (0.4 to 38.4) 42.9 (17.8 to 69.8) 42.9 (17.8 to 69.8)
Georgilis, 1997 17.9 (5.5 to 34.6) 78.6 (61.2 to 92.1) 3.6 (0.0 to 14.7)
Gonzalez-Hevia, 1990 7.1 (0.0 to 28.2) 64.3 (37.1 to 87.6) 28.6 (7.5 to 55.4)
Ispahani, 2000 31.7 (23.1 to 41.0) 43.3 (33.9 to 52.9) 25.0 (17.1 to 33.8)
Katiyo, 2019 28.5 (26.6 to 30.5) 55.5 (53.3 to 57.6) 16.0 (14.4 to 17.6)
Kedzierska, 2008 0.0 (0.0 to 5.7) 86.7 (71.8 to 96.9) 13.3 (3.1 to 28.2)
Koch, 2011 28.4 (23.1 to 34.0) 60.2 (54.2 to 66.1) 11.4 (7.8 to 15.5)
Lester, 1991 25.7 (18.9 to 33.0) 64.2 (56.3 to 71.7) 10.1 (5.7 to 15.6)
Mandal, 1988 55.6 (46.1 to 64.8) 8.3 (3.8 to 14.4) 36.1 (27.3 to 45.4)
Matas, 1995 0.0 (0.0 to 5.3) 100.0 (94.7 to 100.0) 0.0 (0.0 to 5.3)
Noriega, 1994 38.5 (20.5 to 58.1) 61.5 (41.9 to 79.5) 0.0 (0.0 to 6.5)
Papaevangelou, 2004 18.2 (11.5 to 26.0) 66.4 (57.2 to 74.9) 15.5 (9.2 to 22.9)
Preveden, 2001 0.0 (0.0 to 14.0) 83.3 (56.0 to 99.5) 16.7 (0.5 to 44.0)
Prignet, 1993 31.8 (13.8 to 53.0) 45.5 (25.0 to 66.7) 22.7 (7.3 to 42.9)
Ramos, 1996 23.3 (14.2 to 33.7) 76.7 (66.3 to 85.8) 0.0 (0.0 to 2.3)
Yombi, 2015 61.1 (37.3 to 82.6) 38.9 (17.4 to 62.7) 0.0 (0.0 to 9.4)
Pooled 21.4 (12.4 to 29.9) 67.6 (54.2 to 74.8) 11.1 (4.8 to 18.3)
I-squared: 95.3 (93.8-96.4)
Cochran's Q: 404.2
Chi2, p <0.001
tau2: 0.206
0 20 40 60 80 100
Prevalence (%)
0 20 40 60 80 100
Prevalence (%)
0 20 40 60 80 100
Prevalence (%)

Supplemental Figure 8 - Forest plot of meta-analysis of prevalence of non-typhoidal *Salmonella enterica* from normally sterile sites of all serogrouped isolates: serogroup O:4, serogroup O:9, and other serogroups per age groups, 1941-2019 (82 articles, 24,258 isolates)

### Adults

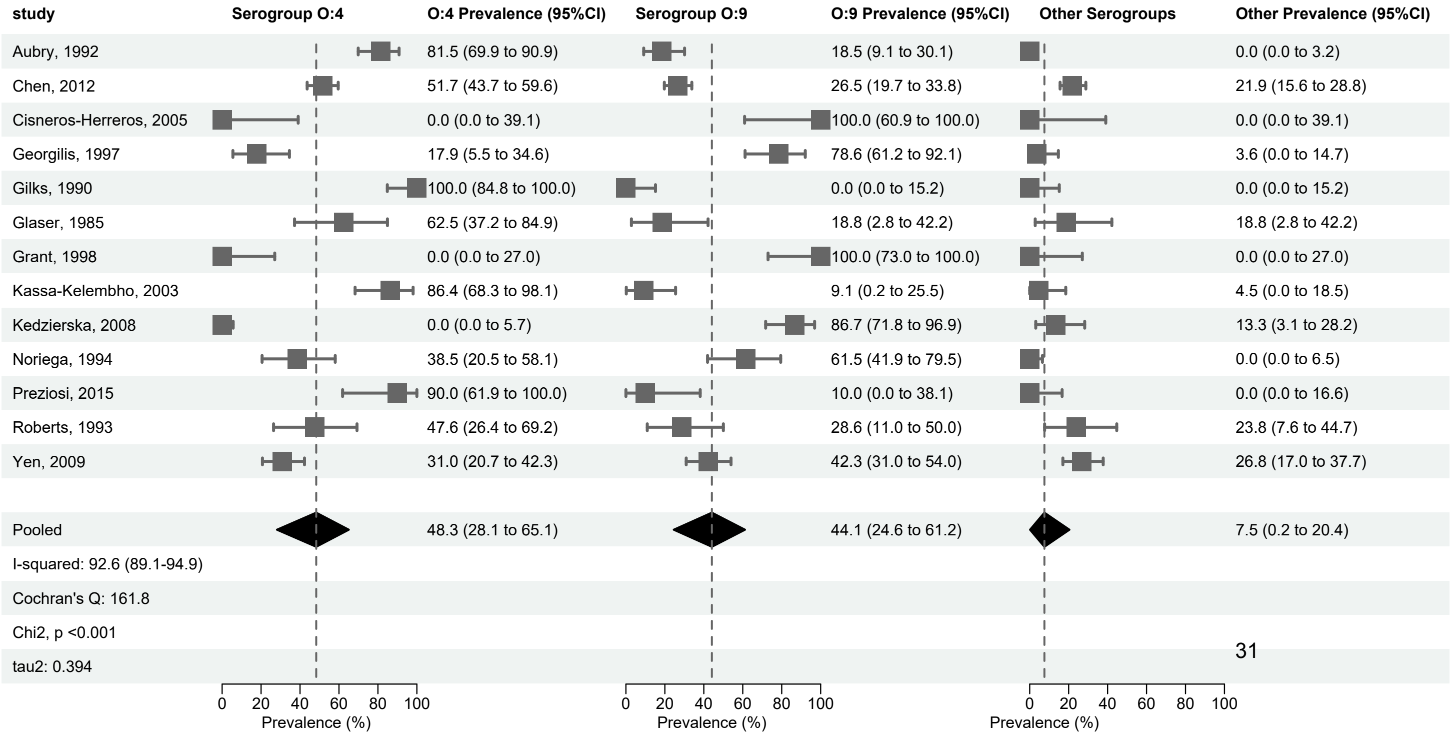

study
Serogroup O:4
O:4 Prevalence (95%CI)
Serogroup O:9
O:9 Prevalence (95%CI)
Other Serogroups
Other Prevalence (95%CI)
Aubry, 1992
81.5 (69.9 to 90.9)
18.5 (9.1 to 30.1)
0.0 (0.0 to 3.2)
Chen, 2012
51.7 (43.7 to 59.6)
26.5 (19.7 to 33.8)
21.9 (15.6 to 28.8)
Cisneros-Herreros, 2005
0.0 (0.0 to 39.1)
100.0 (60.9 to 100.0)
0.0 (0.0 to 39.1)
Georgilis, 1997
17.9 (5.5 to 34.6)
78.6 (61.2 to 92.1)
3.6 (0.0 to 14.7)
Gilks, 1990
100.0 (84.8 to 100.0)
0.0 (0.0 to 15.2)
0.0 (0.0 to 15.2)
Glaser, 1985
62.5 (37.2 to 84.9)
18.8 (2.8 to 42.2)
18.8 (2.8 to 42.2)
Grant, 1998
0.0 (0.0 to 27.0)
100.0 (73.0 to 100.0)
0.0 (0.0 to 27.0)
Kassa-Kelembho, 2003
86.4 (68.3 to 98.1)
9.1 (0.2 to 25.5)
4.5 (0.0 to 18.5)
Kedzierska, 2008
0.0 (0.0 to 5.7)
86.7 (71.8 to 96.9)
13.3 (3.1 to 28.2)
Noriega, 1994
38.5 (20.5 to 58.1)
61.5 (41.9 to 79.5)
0.0 (0.0 to 6.5)
Preziosi, 2015
90.0 (61.9 to 100.0)
10.0 (0.0 to 38.1)
0.0 (0.0 to 16.6)
Roberts, 1993
47.6 (26.4 to 69.2)
28.6 (11.0 to 50.0)
23.8 (7.6 to 44.7)
Yen, 2009
31.0 (20.7 to 42.3)
42.3 (31.0 to 54.0)
26.8 (17.0 to 37.7)
Pooled
48.3 (28.1 to 65.1)
44.1 (24.6 to 61.2)
7.5 (0.2 to 20.4)
I-squared: 92.6 (89.1-94.9)
Cochran's Q: 161.8
Chi2, p <0.001
tau2: 0.394
0
20
40
60
80
100
Prevalence (%)
0
20
40
60
80
100
Prevalence (%)
0
20
40
60
80
100
Prevalence (%)

### Children

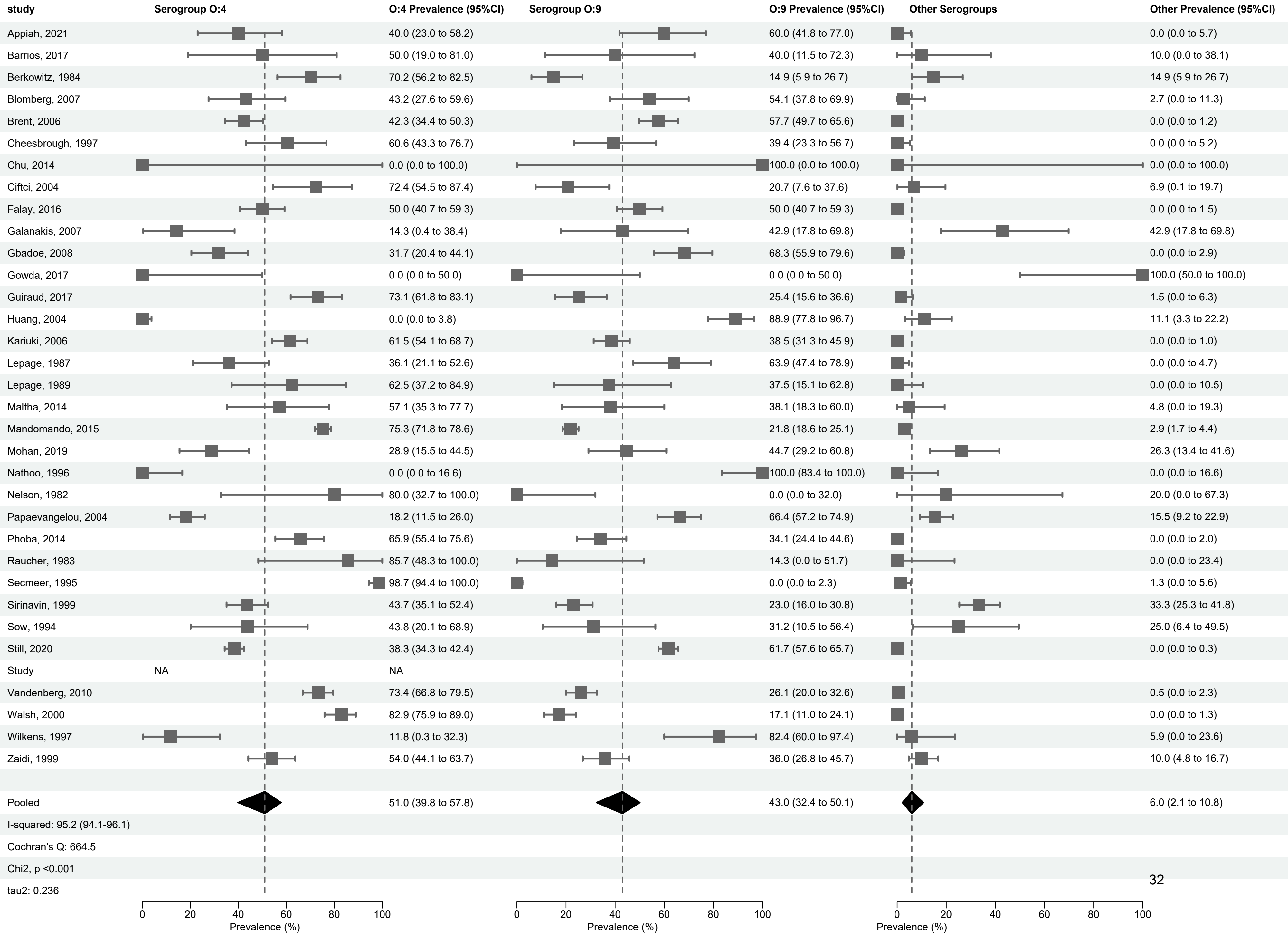
study
Serogroup O:4
O:4 Prevalence (95%CI)
Serogroup O:9
O:9 Prevalence (95%CI)
Other Serogroups
Other Prevalence (95%CI)
Appiah, 2021 40.0 (23.0 to 58.2) 60.0 (41.8 to 77.0) 0.0 (0.0 to 5.7)
Barrios, 2017 50.0 (19.0 to 81.0) 40.0 (11.5 to 72.3) 10.0 (0.0 to 38.1)
Berkowitz, 1984 70.2 (56.2 to 82.5) 14.9 (5.9 to 26.7) 14.9 (5.9 to 26.7)
Blomberg, 2007 43.2 (27.6 to 59.6) 54.1 (37.8 to 69.9) 2.7 (0.0 to 11.3)
Brent, 2006 42.3 (34.4 to 50.3) 57.7 (49.7 to 65.6) 0.0 (0.0 to 1.2)
Cheesbrough, 1997 60.6 (43.3 to 76.7) 39.4 (23.3 to 56.7) 0.0 (0.0 to 5.2)
Chu, 2014 0.0 (0.0 to 100.0) 100.0 (0.0 to 100.0) 0.0 (0.0 to 100.0)
Ciftci, 2004 72.4 (54.5 to 87.4) 20.7 (7.6 to 37.6) 6.9 (0.1 to 19.7)
Falay, 2016 50.0 (40.7 to 59.3) 50.0 (40.7 to 59.3) 0.0 (0.0 to 1.5)
Galanakis, 2007 14.3 (0.4 to 38.4) 42.9 (17.8 to 69.8) 42.9 (17.8 to 69.8)
Gbadoe, 2008 31.7 (20.4 to 44.1) 68.3 (55.9 to 79.6) 0.0 (0.0 to 2.9)
Gowda, 2017 0.0 (0.0 to 50.0) 0.0 (0.0 to 50.0) 100.0 (50.0 to 100.0)
Guiraud, 2017 73.1 (61.8 to 83.1) 25.4 (15.6 to 36.6) 1.5 (0.0 to 6.3)
Huang, 2004 0.0 (0.0 to 3.8) 88.9 (77.8 to 96.7) 11.1 (3.3 to 22.2)
Kariuki, 2006 61.5 (54.1 to 68.7) 38.5 (31.3 to 45.9) 0.0 (0.0 to 1.0)
Lepage, 1987 36.1 (21.1 to 52.6) 63.9 (47.4 to 78.9) 0.0 (0.0 to 4.7)
Lepage, 1989 62.5 (37.2 to 84.9) 37.5 (15.1 to 62.8) 0.0 (0.0 to 10.5)
Maltha, 2014 57.1 (35.3 to 77.7) 38.1 (18.3 to 60.0) 4.8 (0.0 to 19.3)
Mandomando, 2015 75.3 (71.8 to 78.6) 21.8 (18.6 to 25.1) 2.9 (1.7 to 4.4)
Mohan, 2019 28.9 (15.5 to 44.5) 44.7 (29.2 to 60.8) 26.3 (13.4 to 41.6)
Nathoo, 1996 0.0 (0.0 to 16.6) 100.0 (83.4 to 100.0) 0.0 (0.0 to 16.6)
Nelson, 1982 80.0 (32.7 to 100.0) 0.0 (0.0 to 32.0) 20.0 (0.0 to 67.3)
Papaevangelou, 2004 18.2 (11.5 to 26.0) 66.4 (57.2 to 74.9) 15.5 (9.2 to 22.9)
Phoba, 2014 65.9 (55.4 to 75.6) 34.1 (24.4 to 44.6) 0.0 (0.0 to 2.0)
Raucher, 1983 85.7 (48.3 to 100.0) 14.3 (0.0 to 51.7) 0.0 (0.0 to 23.4)
Secmeer, 1995 98.7 (94.4 to 100.0) 0.0 (0.0 to 2.3) 1.3 (0.0 to 5.6)
Sirinavin, 1999 43.7 (35.1 to 52.4) 23.0 (16.0 to 30.8) 33.3 (25.3 to 41.8)
Sow, 1994 43.8 (20.1 to 68.9) 31.2 (10.5 to 56.4) 25.0 (6.4 to 49.5)
Still, 2020 38.3 (34.3 to 42.4) 61.7 (57.6 to 65.7) 0.0 (0.0 to 0.3)
Study NA NA
Vandenberg, 2010 73.4 (66.8 to 79.5) 26.1 (20.0 to 32.6) 0.5 (0.0 to 2.3)
Walsh, 2000 82.9 (75.9 to 89.0) 17.1 (11.0 to 24.1) 0.0 (0.0 to 1.3)
Wilkens, 1997 11.8 (0.3 to 32.3) 82.4 (60.0 to 97.4) 5.9 (0.0 to 23.6)
Zaidi, 1999 54.0 (44.1 to 63.7) 36.0 (26.8 to 45.7) 10.0 (4.8 to 16.7)
Pooled 51.0 (39.8 to 57.8) 43.0 (32.4 to 50.1) 6.0 (2.1 to 10.8)
I-squared: 95.2 (94.1-96.1)
Cochran's Q: 664.5
Chi2, p <0.001
tau2: 0.236
0 20 40 60 80 100
Prevalence (%)
0 20 40 60 80 100
Prevalence (%)
0 20 40 60 80 100
Prevalence (%)

Mixed

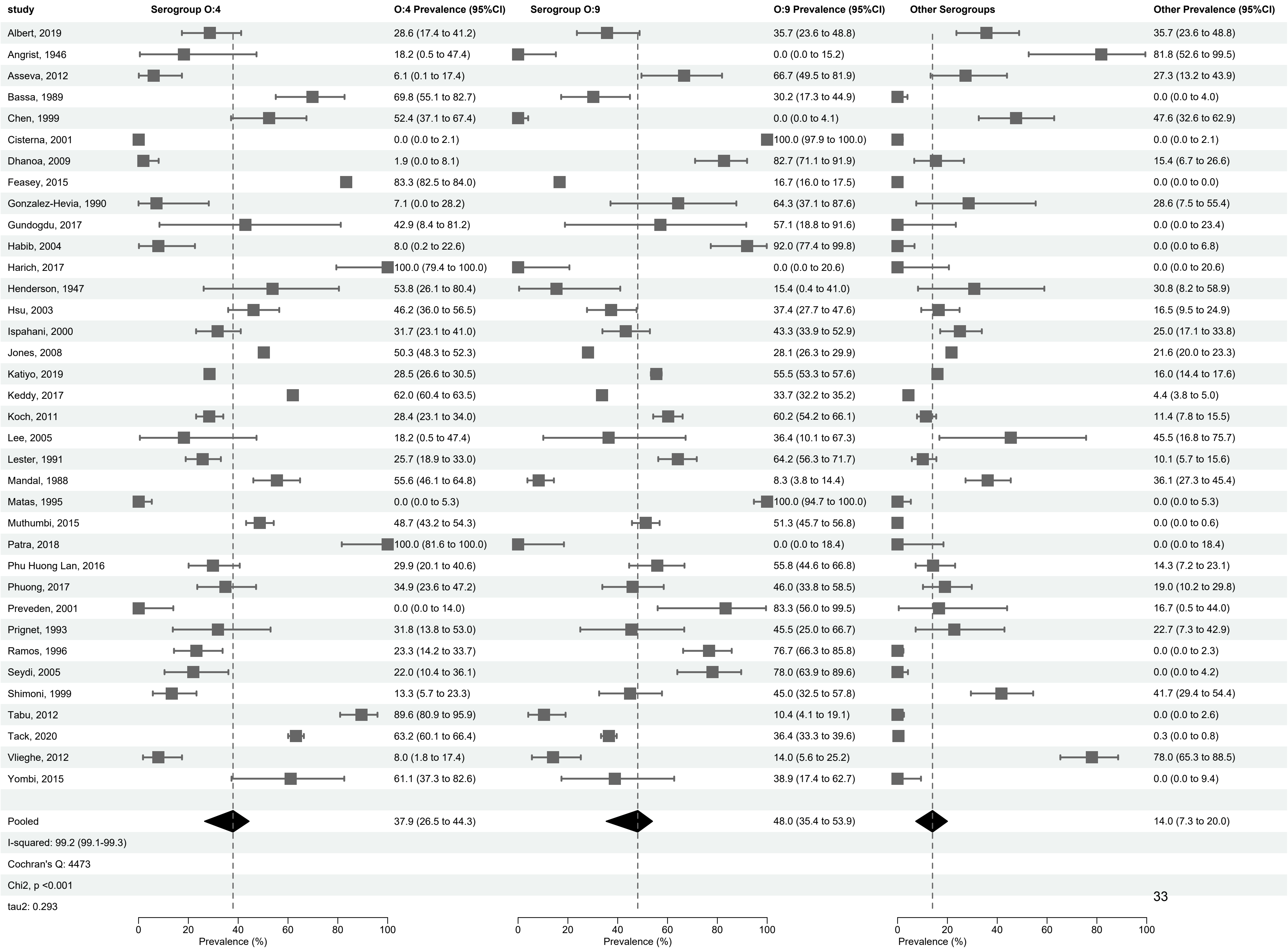
study
Serogroup O:4
O:4 Prevalence (95%CI)
Serogroup O:9
O:9 Prevalence (95%CI)
Other Serogroups
Other Prevalence (95%CI)
Albert, 2019 28.6 (17.4 to 41.2) 35.7 (23.6 to 48.8) 35.7 (23.6 to 48.8)
Angrist, 1946 18.2 (0.5 to 47.4) 0.0 (0.0 to 15.2) 81.8 (52.6 to 99.5)
Asseva, 2012 6.1 (0.1 to 17.4) 66.7 (49.5 to 81.9) 27.3 (13.2 to 43.9)
Bassa, 1989 69.8 (55.1 to 82.7) 30.2 (17.3 to 44.9) 0.0 (0.0 to 4.0)
Chen, 1999 52.4 (37.1 to 67.4) 0.0 (0.0 to 4.1) 47.6 (32.6 to 62.9)
Cisterna, 2001 0.0 (0.0 to 2.1) 100.0 (97.9 to 100.0) 0.0 (0.0 to 2.1)
Dhanoa, 2009 1.9 (0.0 to 8.1) 82.7 (71.1 to 91.9) 15.4 (6.7 to 26.6)
Feasey, 2015 83.3 (82.5 to 84.0) 16.7 (16.0 to 17.5) 0.0 (0.0 to 0.0)
Gonzalez-Hevia, 1990 7.1 (0.0 to 28.2) 64.3 (37.1 to 87.6) 28.6 (7.5 to 55.4)
Gundogdu, 2017 42.9 (8.4 to 81.2) 57.1 (18.8 to 91.6) 0.0 (0.0 to 23.4)
Habib, 2004 8.0 (0.2 to 22.6) 92.0 (77.4 to 99.8) 0.0 (0.0 to 6.8)
Harich, 2017 100.0 (79.4 to 100.0) 0.0 (0.0 to 20.6) 0.0 (0.0 to 20.6)
Henderson, 1947 53.8 (26.1 to 80.4) 15.4 (0.4 to 41.0) 30.8 (8.2 to 58.9)
Hsu, 2003 46.2 (36.0 to 56.5) 37.4 (27.7 to 47.6) 16.5 (9.5 to 24.9)
Ispahani, 2000 31.7 (23.1 to 41.0) 43.3 (33.9 to 52.9) 25.0 (17.1 to 33.8)
Jones, 2008 50.3 (48.3 to 52.3) 28.1 (26.3 to 29.9) 21.6 (20.0 to 23.3)
Katiyo, 2019 28.5 (26.6 to 30.5) 55.5 (53.3 to 57.6) 16.0 (14.4 to 17.6)
Keddy, 2017 62.0 (60.4 to 63.5) 33.7 (32.2 to 35.2) 4.4 (3.8 to 5.0)
Koch, 2011 28.4 (23.1 to 34.0) 60.2 (54.2 to 66.1) 11.4 (7.8 to 15.5)
Lee, 2005 18.2 (0.5 to 47.4) 36.4 (10.1 to 67.3) 45.5 (16.8 to 75.7)
Lester, 1991 25.7 (18.9 to 33.0) 64.2 (56.3 to 71.7) 10.1 (5.7 to 15.6)
Mandal, 1988 55.6 (46.1 to 64.8) 8.3 (3.8 to 14.4) 36.1 (27.3 to 45.4)
Matas, 1995 0.0 (0.0 to 5.3) 100.0 (94.7 to 100.0) 0.0 (0.0 to 5.3)
Muthumbi, 2015 48.7 (43.2 to 54.3) 51.3 (45.7 to 56.8) 0.0 (0.0 to 0.6)
Patra, 2018 100.0 (81.6 to 100.0) 0.0 (0.0 to 18.4) 0.0 (0.0 to 18.4)
Phu Huong Lan, 2016 29.9 (20.1 to 40.6) 55.8 (44.6 to 66.8) 14.3 (7.2 to 23.1)
Phuong, 2017 34.9 (23.6 to 47.2) 46.0 (33.8 to 58.5) 19.0 (10.2 to 29.8)
Preveden, 2001 0.0 (0.0 to 14.0) 83.3 (56.0 to 99.5) 16.7 (0.5 to 44.0)
Prignet, 1993 31.8 (13.8 to 53.0) 45.5 (25.0 to 66.7) 22.7 (7.3 to 42.9)
Ramos, 1996 23.3 (14.2 to 33.7) 76.7 (66.3 to 85.8) 0.0 (0.0 to 2.3)
Seydi, 2005 22.0 (10.4 to 36.1) 78.0 (63.9 to 89.6) 0.0 (0.0 to 4.2)
Shimoni, 1999 13.3 (5.7 to 23.3) 45.0 (32.5 to 57.8) 41.7 (29.4 to 54.4)
Tabu, 2012 89.6 (80.9 to 95.9) 10.4 (4.1 to 19.1) 0.0 (0.0 to 2.6)
Tack, 2020 63.2 (60.1 to 66.4) 36.4 (33.3 to 39.6) 0.3 (0.0 to 0.8)
Vlieghe, 2012 8.0 (1.8 to 17.4) 14.0 (5.6 to 25.2) 78.0 (65.3 to 88.5)
Yombi, 2015 61.1 (37.3 to 82.6) 38.9 (17.4 to 62.7) 0.0 (0.0 to 9.4)
Pooled 37.9 (26.5 to 44.3) 48.0 (35.4 to 53.9) 14.0 (7.3 to 20.0)
I-squared: 99.2 (99.1-99.3)
Cochran's Q: 4473
Chi2, p <0.001
tau2: 0.293
0 20 40 60 80 100
Prevalence (%)
0 20 40 60 80 100
Prevalence (%)
0 20 40 60 80 100
Prevalence (%)

Supplemental Figure 9 - Forest plot of meta-analysis of prevalence of non-typhoidal *Salmonella enterica* serovar Typhimurium, *Salmonella enterica* serovar Enteritidis and other serovars from normally sterile sites, 1941-2019 (79 articles, 23,976 isolates)

#### Serovars

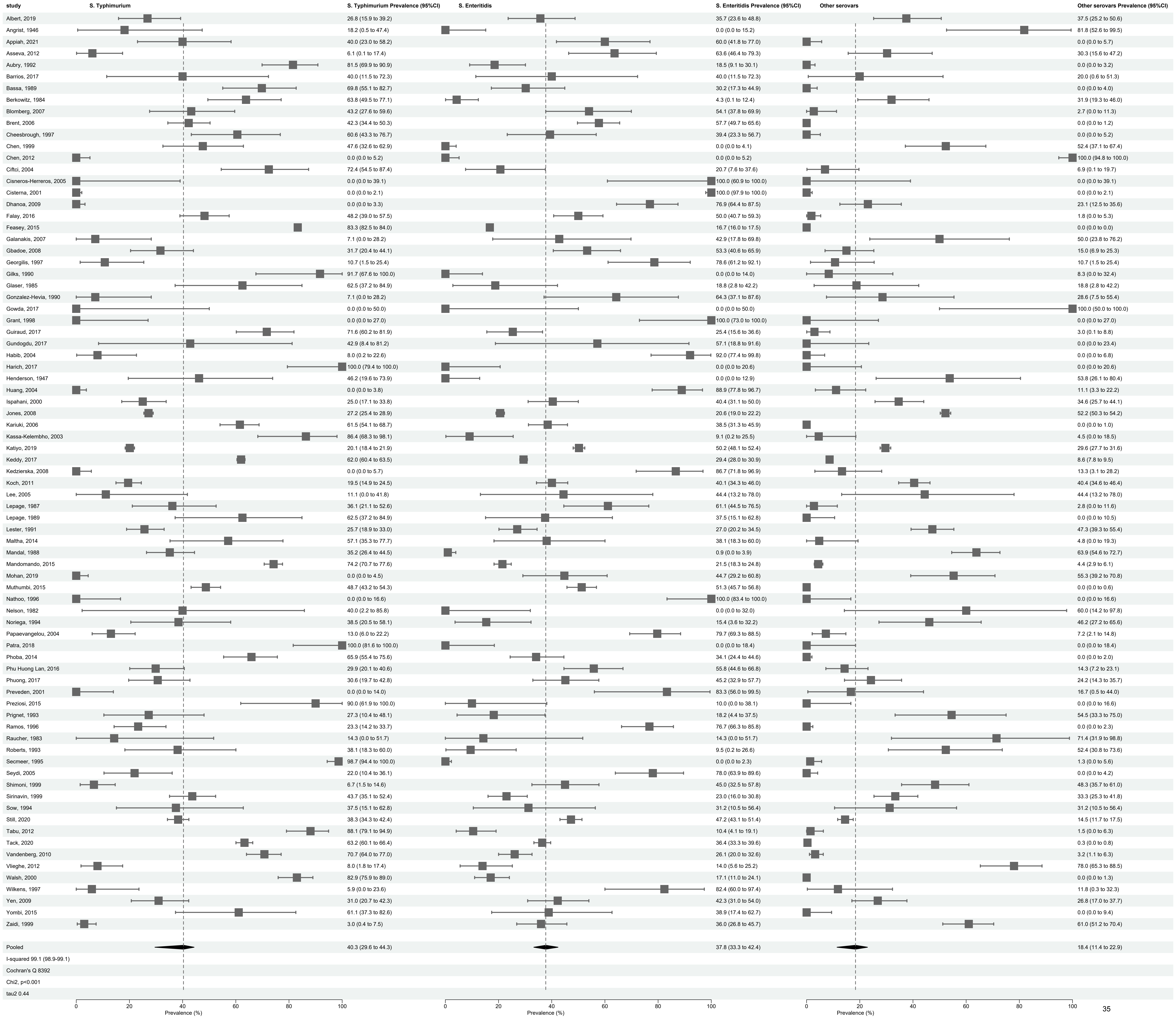


| study | S. Typhimurium Prevalence (95%CI) | S. Enteritidis Prevalence (95%CI) | Other serovars Prevalence (95%CI) |
|---|---|---|---|
| Albert, 2019 | 26.8 (15.9 to 39.2) | 35.7 (23.6 to 48.8) | 37.5 (25.2 to 50.6) |
| Angrist, 1946 | 18.2 (0.5 to 47.4) | 0.0 (0.0 to 15.2) | 81.8 (52.6 to 99.5) |
| Appiah, 2021 | 40.0 (23.0 to 58.2) | 60.0 (41.8 to 77.0) | 0.0 (0.0 to 5.7) |
| Asseva, 2012 | 6.1 (0.1 to 17.4) | 63.6 (46.4 to 79.3) | 30.3 (15.6 to 47.2) |
| Aubry, 1992 | 81.5 (69.9 to 90.9) | 18.5 (9.1 to 30.1) | 0.0 (0.0 to 3.2) |
| Barrios, 2017 | 40.0 (11.5 to 72.3) | 40.0 (11.5 to 72.3) | 20.0 (0.6 to 51.3) |
| Bassa, 1989 | 69.8 (55.1 to 82.7) | 30.2 (17.3 to 44.9) | 0.0 (0.0 to 4.0) |
| Berkowitz, 1984 | 63.8 (49.5 to 77.1) | 4.3 (0.1 to 12.4) | 31.9 (19.3 to 46.0) |
| Blomberg, 2007 | 43.2 (27.6 to 59.6) | 54.1 (37.8 to 69.9) | 2.7 (0.0 to 11.3) |
| Brent, 2006 | 42.3 (34.4 to 50.3) | 57.7 (49.7 to 65.6) | 0.0 (0.0 to 1.2) |
| Cheesbrough, 1997 | 60.6 (43.3 to 76.7) | 39.4 (23.3 to 56.7) | 0.0 (0.0 to 5.2) |
| Chen, 1999 | 47.6 (32.6 to 62.9) | 0.0 (0.0 to 4.1) | 52.4 (37.1 to 67.4) |
| Chen, 2012 | 0.0 (0.0 to 5.2) | 0.0 (0.0 to 5.2) | 100.0 (94.8 to 100.0) |
| Ciftci, 2004 | 72.4 (54.5 to 87.4) | 20.7 (7.6 to 37.6) | 6.9 (0.1 to 19.7) |
| Cisneros-Herreros, 2005 | 0.0 (0.0 to 39.1) | 100.0 (60.9 to 100.0) | 0.0 (0.0 to 39.1) |
| Cisterna, 2001 | 0.0 (0.0 to 2.1) | 100.0 (97.9 to 100.0) | 0.0 (0.0 to 2.1) |
| Dhanoa, 2009 | 0.0 (0.0 to 3.3) | 76.9 (64.4 to 87.5) | 23.1 (12.5 to 35.6) |
| Falay, 2016 | 48.2 (39.0 to 57.5) | 50.0 (40.7 to 59.3) | 1.8 (0.0 to 5.3) |
| Feasey, 2015 | 83.3 (82.5 to 84.0) | 16.7 (16.0 to 17.5) | 0.0 (0.0 to 0.0) |
| Galanakis, 2007 | 7.1 (0.0 to 28.2) | 42.9 (17.8 to 69.8) | 50.0 (23.8 to 76.2) |
| Gbadoe, 2008 | 31.7 (20.4 to 44.1) | 53.3 (40.6 to 65.9) | 15.0 (6.9 to 25.3) |
| Georgilis, 1997 | 10.7 (1.5 to 25.4) | 78.6 (61.2 to 92.1) | 10.7 (1.5 to 25.4) |
| Gilks, 1990 | 91.7 (67.6 to 100.0) | 0.0 (0.0 to 14.0) | 8.3 (0.0 to 32.4) |
| Glaser, 1985 | 62.5 (37.2 to 84.9) | 18.8 (2.8 to 42.2) | 18.8 (2.8 to 42.2) |
| Gonzalez-Hevia, 1990 | 7.1 (0.0 to 28.2) | 64.3 (37.1 to 87.6) | 28.6 (7.5 to 55.4) |
| Gowda, 2017 | 0.0 (0.0 to 50.0) | 0.0 (0.0 to 50.0) | 100.0 (50.0 to 100.0) |
| Grant, 1998 | 0.0 (0.0 to 27.0) | 100.0 (73.0 to 100.0) | 0.0 (0.0 to 27.0) |
| Guiraud, 2017 | 71.6 (60.2 to 81.9) | 25.4 (15.6 to 36.6) | 3.0 (0.1 to 8.8) |
| Gundogdu, 2017 | 42.9 (8.4 to 81.2) | 57.1 (18.8 to 91.6) | 0.0 (0.0 to 23.4) |
| Habib, 2004 | 8.0 (0.2 to 22.6) | 92.0 (77.4 to 99.8) | 0.0 (0.0 to 6.8) |
| Harich, 2017 | 100.0 (79.4 to 100.0) | 0.0 (0.0 to 20.6) | 0.0 (0.0 to 20.6) |
| Henderson, 1947 | 46.2 (19.6 to 73.9) | 0.0 (0.0 to 12.9) | 53.8 (26.1 to 80.4) |
| Huang, 2004 | 0.0 (0.0 to 3.8) | 88.9 (77.8 to 96.7) | 11.1 (3.3 to 22.2) |
| Ispahani, 2000 | 25.0 (17.1 to 33.8) | 40.4 (31.1 to 50.0) | 34.6 (25.7 to 44.1) |
| Jones, 2008 | 27.2 (25.4 to 28.9) | 20.6 (19.0 to 22.2) | 52.2 (50.3 to 54.2) |
| Kariuki, 2006 | 61.5 (54.1 to 68.7) | 38.5 (31.3 to 45.9) | 0.0 (0.0 to 1.0) |
| Kassa-Kelembho, 2003 | 86.4 (68.3 to 98.1) | 9.1 (0.2 to 25.5) | 4.5 (0.0 to 18.5) |
| Katiyo, 2019 | 20.1 (18.4 to 21.9) | 50.2 (48.1 to 52.4) | 29.6 (27.7 to 31.6) |
| Keddy, 2017 | 62.0 (60.4 to 63.5) | 29.4 (28.0 to 30.9) | 8.6 (7.8 to 9.5) |
| Kedzierska, 2008 | 0.0 (0.0 to 5.7) | 86.7 (71.8 to 96.9) | 13.3 (3.1 to 28.2) |
| Koch, 2011 | 19.5 (14.9 to 24.5) | 40.1 (34.3 to 46.0) | 40.4 (34.6 to 46.4) |
| Lee, 2005 | 11.1 (0.0 to 41.8) | 44.4 (13.2 to 78.0) | 44.4 (13.2 to 78.0) |
| Lepage, 1987 | 36.1 (21.1 to 52.6) | 61.1 (44.5 to 76.5) | 2.8 (0.0 to 11.6) |
| Lepage, 1989 | 62.5 (37.2 to 84.9) | 37.5 (15.1 to 62.8) | 0.0 (0.0 to 10.5) |
| Lester, 1991 | 25.7 (18.9 to 33.0) | 27.0 (20.2 to 34.5) | 47.3 (39.3 to 55.4) |
| Maltha, 2014 | 57.1 (35.3 to 77.7) | 38.1 (18.3 to 60.0) | 4.8 (0.0 to 19.3) |
| Mandal, 1988 | 35.2 (26.4 to 44.5) | 0.9 (0.0 to 3.9) | 63.9 (54.6 to 72.7) |
| Mandomando, 2015 | 74.2 (70.7 to 77.6) | 21.5 (18.3 to 24.8) | 4.4 (2.9 to 6.1) |
| Mohan, 2019 | 0.0 (0.0 to 4.5) | 44.7 (29.2 to 60.8) | 55.3 (39.2 to 70.8) |
| Muthumbi, 2015 | 48.7 (43.2 to 54.3) | 51.3 (45.7 to 56.8) | 0.0 (0.0 to 0.6) |
| Nathoo, 1996 | 0.0 (0.0 to 16.6) | 100.0 (83.4 to 100.0) | 0.0 (0.0 to 16.6) |
| Nelson, 1982 | 40.0 (2.2 to 85.8) | 0.0 (0.0 to 32.0) | 60.0 (14.2 to 97.8) |
| Noriega, 1994 | 38.5 (20.5 to 58.1) | 15.4 (3.6 to 32.2) | 46.2 (27.2 to 65.6) |
| Papaevangelou, 2004 | 13.0 (6.0 to 22.2) | 79.7 (69.3 to 88.5) | 7.2 (2.1 to 14.8) |
| Patra, 2018 | 100.0 (81.6 to 100.0) | 0.0 (0.0 to 18.4) | 0.0 (0.0 to 18.4) |
| Phoba, 2014 | 65.9 (55.4 to 75.6) | 34.1 (24.4 to 44.6) | 0.0 (0.0 to 2.0) |
| Phu Huong Lan, 2016 | 29.9 (20.1 to 40.6) | 55.8 (44.6 to 66.8) | 14.3 (7.2 to 23.1) |
| Phuong, 2017 | 30.6 (19.7 to 42.8) | 45.2 (32.9 to 57.7) | 24.2 (14.3 to 35.7) |
| Preveden, 2001 | 0.0 (0.0 to 14.0) | 83.3 (56.0 to 99.5) | 16.7 (0.5 to 44.0) |
| Preziosi, 2015 | 90.0 (61.9 to 100.0) | 10.0 (0.0 to 38.1) | 0.0 (0.0 to 16.6) |
| Prignet, 1993 | 27.3 (10.4 to 48.1) | 18.2 (4.4 to 37.5) | 54.5 (33.3 to 75.0) |
| Ramos, 1996 | 23.3 (14.2 to 33.7) | 76.7 (66.3 to 85.8) | 0.0 (0.0 to 2.3) |
| Raucher, 1983 | 14.3 (0.0 to 51.7) | 14.3 (0.0 to 51.7) | 71.4 (31.9 to 98.8) |
| Roberts, 1993 | 38.1 (18.3 to 60.0) | 9.5 (0.2 to 26.6) | 52.4 (30.8 to 73.6) |
| Secmeer, 1995 | 98.7 (94.4 to 100.0) | 0.0 (0.0 to 2.3) | 1.3 (0.0 to 5.6) |
| Seydi, 2005 | 22.0 (10.4 to 36.1) | 78.0 (63.9 to 89.6) | 0.0 (0.0 to 4.2) |
| Shimoni, 1999 | 6.7 (1.5 to 14.6) | 45.0 (32.5 to 57.8) | 48.3 (35.7 to 61.0) |
| Sirinavin, 1999 | 43.7 (35.1 to 52.4) | 23.0 (16.0 to 30.8) | 33.3 (25.3 to 41.8) |
| Sow, 1994 | 37.5 (15.1 to 62.8) | 31.2 (10.5 to 56.4) | 31.2 (10.5 to 56.4) |
| Still, 2020 | 38.3 (34.3 to 42.4) | 47.2 (43.1 to 51.4) | 14.5 (11.7 to 17.5) |
| Tabu, 2012 | 88.1 (79.1 to 94.9) | 10.4 (4.1 to 19.1) | 1.5 (0.0 to 6.3) |
| Tack, 2020 | 63.2 (60.1 to 66.4) | 36.4 (33.3 to 39.6) | 0.3 (0.0 to 0.8) |
| Vandenberg, 2010 | 70.7 (64.0 to 77.0) | 26.1 (20.0 to 32.6) | 3.2 (1.1 to 6.3) |
| Vlieghe, 2012 | 8.0 (1.8 to 17.4) | 14.0 (5.6 to 25.2) | 78.0 (65.3 to 88.5) |
| Walsh, 2000 | 82.9 (75.9 to 89.0) | 17.1 (11.0 to 24.1) | 0.0 (0.0 to 1.3) |
| Wilkens, 1997 | 5.9 (0.0 to 23.6) | 82.4 (60.0 to 97.4) | 11.8 (0.3 to 32.3) |
| Yen, 2009 | 31.0 (20.7 to 42.3) | 42.3 (31.0 to 54.0) | 26.8 (17.0 to 37.7) |
| Yombi, 2015 | 61.1 (37.3 to 82.6) | 38.9 (17.4 to 62.7) | 0.0 (0.0 to 9.4) |
| Zaidi, 1999 | 3.0 (0.4 to 7.5) | 36.0 (26.8 to 45.7) | 61.0 (51.2 to 70.4) |
| Pooled | 40.3 (29.6 to 44.3) | 37.8 (33.3 to 42.4) | 18.4 (11.4 to 22.9) |
| I-squared 99.1 (98.9-99.1) | | | |
| Cochran's Q 8392 | | | |
| Chi2, p<0.001 | | | |
| tau2 0.44 | | | |
